# Targeting Polθ Helicase with RP-3467: Preclinical Validation and Clinical Response in a g*BRCA1* ovarian cancer patient with an acquired PARPi resistance mutation in *TP53BP1*

**DOI:** 10.64898/2026.09.16.26363059

**Authors:** Ezra Y. Rosen, Marie-Claude Mathieu, Gino B. Ferraro, Philippe Mochirian, Claude Godbout, Hugo Poirier, Sara Fournier, Danielle Henry, Hyeyeon Kim, Robert Papp, Shou Yun Yin, Marie-Eve Leclaire, Robert Houle, Parham Nejad, Joseph D. Schonhoft, Karl E. Zahn, W. Cameron Black, Michel Gallant, Insil Kim, Danielle Ulanet, Paul Basciano, Artur Veloso, Victoria Rimkunas, Timothy A. Yap, Robin Guo, Nicholas Mai, Maria Koehler, Michael Zinda, Anne Roulston, Michal Zimmermann, Stephen J. Morris, Agnel Sfeir

**Affiliations:** Department of Medicine, Memorial Sloan Kettering Cancer Center, New York, NY, USA; Repare Therapeutics Inc., Saint Laurent, Quebec, Canada; Repare Therapeutics Inc., Cambridge, MA, USA; Investigational Cancer Therapeutics (Phase I Program), The University of Texas MD Anderson Cancer Center, Houston, TX, USA; Molecular Biology Program, Sloan Kettering Institute, Memorial Sloan Kettering Cancer Center, New York, NY, USA

**Keywords:** Microhomology-mediated end-joining (MMEJ), polymerase theta (Polθ), mitotic repair, DNA double-strand breaks (DSBs), homologous recombination deficient (HRD), BRCA1, BRCA2, ovarian cancer

## Abstract

Homologous recombination (HR) deficiency underlies a range of cancers, and PARP inhibitors (PARPi) targeting this vulnerability have demonstrated clinical efficacy in *BRCA1/2*-mutant tumors. However, resistance mechanisms and tolerability issues limit their long-term clinical effectiveness. Here, we characterize RP-3467, a novel, selective small-molecule inhibitor targeting the ATPase domain of DNA polymerase theta (Polθ), the key mediator of the error-prone microhomology-mediated end joining (MMEJ) pathway. We demonstrate that RP-3467 potently suppresses Polθ enzymatic activity, abrogates MMEJ, and disrupts mitotic DNA repair in HR-deficient (HRD) models, inducing synthetic lethality. In preclinical studies, we show that RP-3467 synergizes strongly with PARPi (olaparib and rucaparib), achieving sustained tumor regression across multiple xenograft and patient-derived xenograft (PDX) models without added systemic toxicity. Mechanistically, we find that RP-3467 increases mitotic DNA damage in vivo, as evidenced by elevated micronuclei and CIP2A foci. RP-3467 was evaluated in a phase 1 clinical trial (NCT06560632) and has shown preliminary signs of clinical activity. A heavily pretreated germline *BRCA1*-mutant ovarian cancer patient achieved a confirmed partial response to RP-3467 plus olaparib (RECIST v1.1), with retrospective profiling revealing a *TP53BP1* loss-of-function mutation previously linked to both PARPi resistance and POLQ dependency. Although broader clinical validation is needed, these early findings suggest that Polθ inhibition has the potential to address PARPi resistance and offer therapeutic benefit in HRD cancers.

## INTRODUCTION

DNA double-strand breaks (DSBs) are among the most lethal forms of genomic damage, arising from both endogenous sources, such as replication fork collapse, and exogenous agents, including ionizing radiation and genotoxic chemotherapy. Accurate DSB repair is essential for maintaining genomic stability and ensuring cell survival. In mammalian cells, DSBs are primarily resolved by two canonical pathways: homologous recombination (HR), a high-fidelity, template-directed mechanism active during S and G2 phases of the cell cycle^1^, and non-homologous end joining (NHEJ), a faster, though non-templated, pathway that operates throughout the cell cycle^2^. In addition to these, a distinct and inherently mutagenic mechanism known as microhomology-mediated end joining (MMEJ) has been identified. This latter pathway largely depends on DNA polymerase theta (Polθ) and serves as a critical backup when HR and NHEJ are compromised^3^.

A landmark example of exploiting DNA repair deficiencies for therapeutic benefit is the development of poly(ADP-ribose) polymerase inhibitors (PARPi) for HR-deficient (HRD) cancers with mutations in *BRCA1*, *BRCA2*, and *PALB2*^4–6^. These agents act by inhibiting PARP1/2 enzymatic activity and blocking single-strand break repair, increasing the accumulation of single-stranded DNA gaps, and trapping PARP enzymes on DNA, ultimately generating replication-associated DSB^7–9^. In HRD cells, the inability to repair these breaks results in synthetic lethality. The clinical utility of PARPi is now firmly established, with approvals in multiple settings as monotherapy or in combination with other agents for *BRCA1/2*-mutated and/or HRD breast, ovarian, prostate, and pancreatic cancers. Ongoing clinical trials are expanding the application of PARPi through rational combination strategies with DNA damage response (DDR) inhibitors or immunotherapies^13^.

Despite this progress, the emergence of acquired resistance remains an obstacle to maximizing the clinical benefit of PARPi^14^. Multiple distinct resistance mechanisms have been described in preclinical models, including HR reactivation through BRCA1/2 reversion mutations or loss of the 53BP1/Shieldin pathway, enhanced replication fork stability, increased drug efflux and *PARP1* mutations^14–16^. While *BRCA1/2* reversions remain the most frequently detected mutations in the clinic, longitudinal sequencing studies of PARPi-treated tumors are revealing the emergence of additional resistance mechanisms, including deleterious mutations in *TP53BP1*^17,18^. Although combination strategies involving DDR inhibitors designed to overcome acquired resistance – such as combinations with inhibitors of ATR, CHK1, or WEE1 – have shown promising synergy in preclinical studies, their clinical utility has been limited by narrow therapeutic windows and overlapping toxicities, including bone marrow suppression and gastrointestinal side effects^13,19–22^.

An emerging therapeutic strategy to enhance the efficacy of PARP inhibitors (PARPi) and potentially delay or overcome resistance involves targeting Polθ. In addition to functioning as a backup repair pathway to resolve DSBs and replication-associated single-stranded gaps in interphase, MMEJ is the dominant DSB repair pathway in mitosis, where it is essential for resolving DNA breaks that persist from S phase, including those caused by replication stress or PARPi-induced fork collapse^23–25^. Polθ is structurally unique, harboring both an N-terminal helicase-like ATPase domain and a C-terminal A-family polymerase domain, each essential for MMEJ activity^26^. Notably, Polθ is minimally expressed in most normal tissues but frequently overexpressed in cancers, especially those with HRD, making it a compelling and selective therapeutic target^27^. While MMEJ is intrinsically mutagenic, it becomes a critical survival mechanism in HRD cells, where high-fidelity repair is compromised, rendering Polθ activity indispensable^28,29^. Moreover, MMEJ plays a compensatory role in cells deficient in key NHEJ factors such as 53BP1^30–32^. These findings have spurred the development of small-molecule inhibitors targeting its polymerase activity, including ART558, RP-6685, and RTx-161, all of which exhibit potent activity in HRD models, especially in combination with PARPi^32–34^.

More recently, the helicase-like ATPase domain of Polθ has been recognized as a mechanistically distinct and druggable vulnerability. The antibiotic novobiocin has been shown in preclinical studies to inhibit Polθ ATPase activity and has since advanced into early-phase clinical trials for HRD tumors^35^. This domain functions mechanistically upstream of DNA synthesis by displacing replication protein A (RPA) from resected DSB ends and, independently, facilitating the annealing of short microhomologies to initiate MMEJ repair^36–38^. Here, we report the discovery and preclinical validation of RP-3467, a small-molecule inhibitor of the Polθ helicase domain. RP-3467 blocks ATPase activity and suppresses MMEJ, leading to synthetic lethality in HRD models. In HRD xenograft models, RP-3467 demonstrates excellent tolerability and robust antitumor activity in combination with PARPi. Early clinical data from an ongoing Phase 1 trial show tumor regression and a molecular response in a patient with PARPi pre-treated *BRCA1*-mutant ovarian cancer with a deleterious mutation in *TP53BP1*, thus providing the initial clinical proof-of-concept for Polθ helicase inhibition as a viable therapeutic strategy.

## RESULTS

### Discovery and characterization of RP-3467 as a selective Polθ inhibitor

We identified RP-3467 as a potent and selective inhibitor of the ATPase activity of Polθ through iterative structure-activity optimization of a lead compound RP-2119^58^ (**Figure 1a**). In a biochemical assay (ADP-glo) that measures ATP consumption by a recombinant His-SUMO-tagged Polθ ATPase domain (amino acids 1-894), RP-3467 showed sub-nanomolar potency (**Figure 1b**), while displaying no discernible inhibitory activity against other DNA repair-related ATPases WRN, BLM, and FANCM (**Extended Data Figure 1a**). Cellular target engagement was confirmed using a Cellular Thermal Shift Assay (CETSA), in which RP-3467 stabilized an ectopically expressed Polθ ATPase construct at single-digit nanomolar concentrations (**Figure 1c**). To assess the functional inhibition of endogenous Polθ activity, we employed a CRISPR/Cas9-based MMEJ repair assay in HCT116 cells. A targeted DSB was induced at the *AAVS1* locus, and repair outcomes were quantified via sequencing-based inference of CRISPR edits (ICE) (**Figure 1d, Extended Data Figure 1b-d**). RP-3467 suppressed MMEJ repair events with an IC_50_ of 3 ± 2 nM, in line with the concentration range showing cellular target engagement (**Figure 1e**).

**Figure 1:**
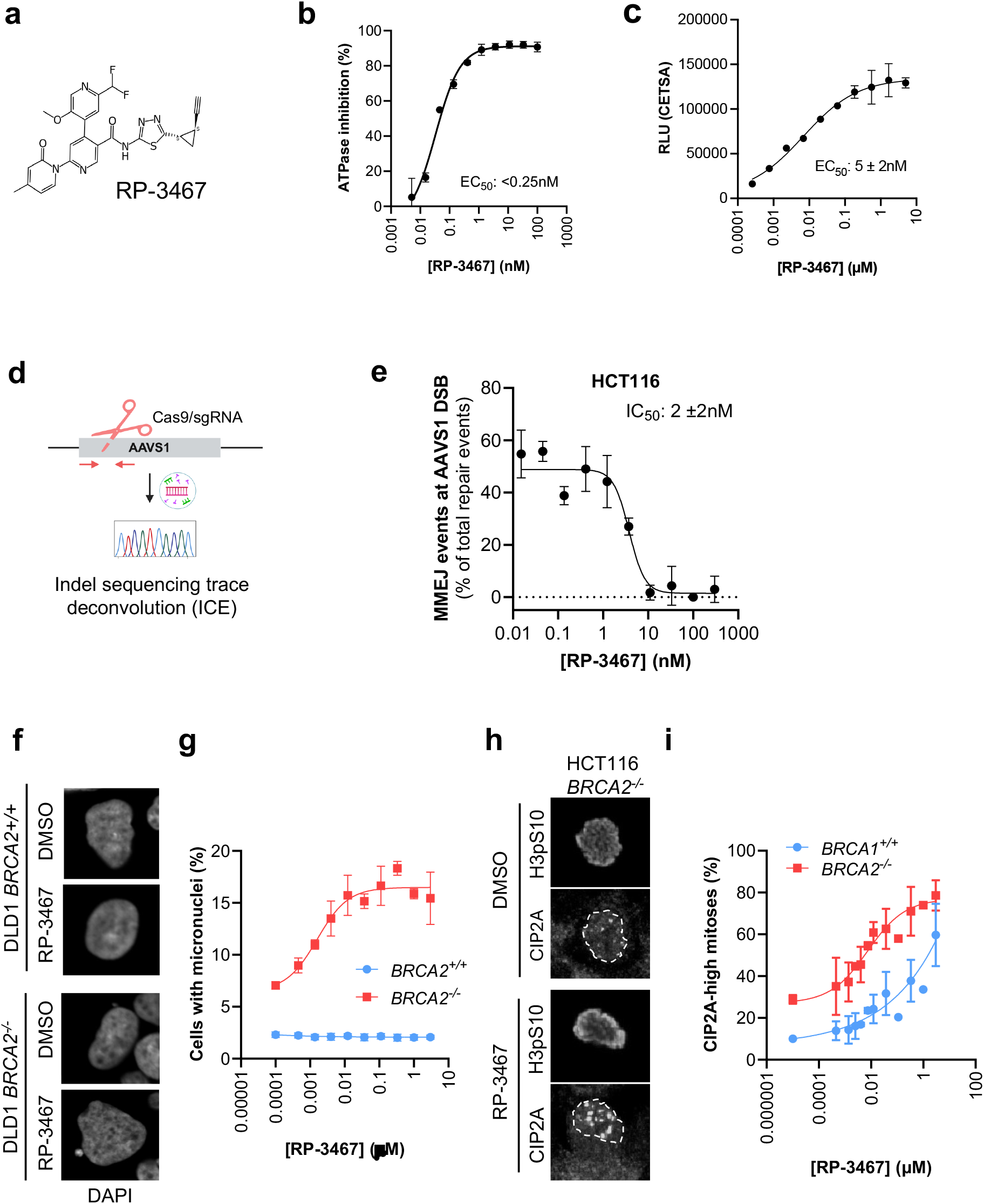
RP-3467 inhibits Polθ ATPase activity and disrupts MMEJ and mitotic DNA repair in HRD cells. **a.** Structure of RP-3467. **b.** Representative ADPglo dose-response assay measuring inhibition of recombinant Polθ ATPase by RP-3467. **I**C_50_ value is a mean of N=5 independent experiments. **c.** Representative CETSA dose-response plot showing thermal stabilization of overexpressed Polθ(1-894)-ePLC in K562 cells. EC_50_ value is a mean of N=6 independent experiments ±SD. **d.** Schematic of a cellular MMEJ assay measuring the repair of a CRISPR/Cas9-induced DNA DSB in the *AAVS1* locus of HCT116 cells. See Methods for details. **e,** Representative RP-3467 dose-response in a MMEJ assay as in d. IC_50_ value is a mean of N=2 independent experiments ±SD. **f-i.** Inhibition of mitotic DNA repair by RP-3467. **f.** Example images of DAPI-stained DLD1 *BRCA2^+/+^* and *BRCA2^-/-^* cells with or without RP-3467 treatment. A micronucleus is highlighted with an arrow. **g.** Quantification of micronuclei-containing cells after indicated RP-3467 concentrations in DLD1 *BRCA2^+/+^* and *BRCA2^-/-^*cells. **h.** Example images of HCT116 *BRCA2^-/-^* cells processed for immunofluorescence with antibodies against phosphorylated histone H3 (H3pS10, mitotic marker) and CIP2A (mitotic DNA damage). **i.** Quantification of CIP2A-high (with nuclear CIP2A intensity greater than the 90th percentile in untreated HCT116 *BRCA2^+/+^* cells) mitotic HCT116 *BRCA2^+/+^* and *BRCA2^-/-^* cells after treatment with indicated RP-3467 concentrations. Data in g and i are mean of N=3 independent experiments ±SD.

Given that Polθ activity has been implicated in mitotic DNA repair, particularly in HRD contexts^24^, we next evaluated the impact of RP-3467 on mitotic DNA damage. Consistent with prior reports linking Polθ loss to increased micronuclei formation in *BRCA2*-deficient cells^39,40^, RP-3467 treatment led to a dose-dependent accumulation of micronuclei in *BRCA2^-/-^* DLD1 cells but not in their *BRCA2^+/+^* counterparts (**Figure 1f,g**). In addition, we examined CIP2A, a marker of unresolved mitotic DNA damage^41,42^, and observed a concentration-dependent increase in CIP2A-positive mitoses following RP-3467 treatment in both *BRCA2*-proficient and -deficient HCT116 cells (**Figure 1h,i**). The concentrations at which these phenotypes emerged were consistent with those required for Polθ target engagement and MMEJ suppression. In conclusion, our results demonstrate that RP-3467 is a selective and potent inhibitor of Polθ ATPase function, suppressing MMEJ repair and inducing mitotic DNA damage phenotypes in HRD settings.

### RP-3467 induces synthetic lethality in HR-deficient cells and synergizes with PARP inhibition

To evaluate the cellular activity of RP-3467, we performed Incucyte-based cell proliferation assays in isogenic *BRCA2*-deficient and -proficient colorectal cancer cell lines (HCT116 and DLD1), as well as in DOTC24510, a high-grade serous ovarian carcinoma cell line harboring a *BRCA2* truncating mutation (p.R3128*). RP-3467 elicited potent, dose-dependent growth inhibition in *BRCA2*-deficient HCT116 and DLD1 cells, with IC_50_ values of 7 ± 3 nM and 4 ± 1 nM, respectively, while showing no measurable effect in their wild-type counterparts (IC_50_ >15 µM; **Figure 2 a,b**). Similar sensitivity was observed in DOTC24510 cells, with an IC_50_ of 15 ± 5 nM (**Figure 2c**). These findings confirm the selective antiproliferative activity of RP-3467 in HR-deficient cellular contexts.

**Figure 2:**
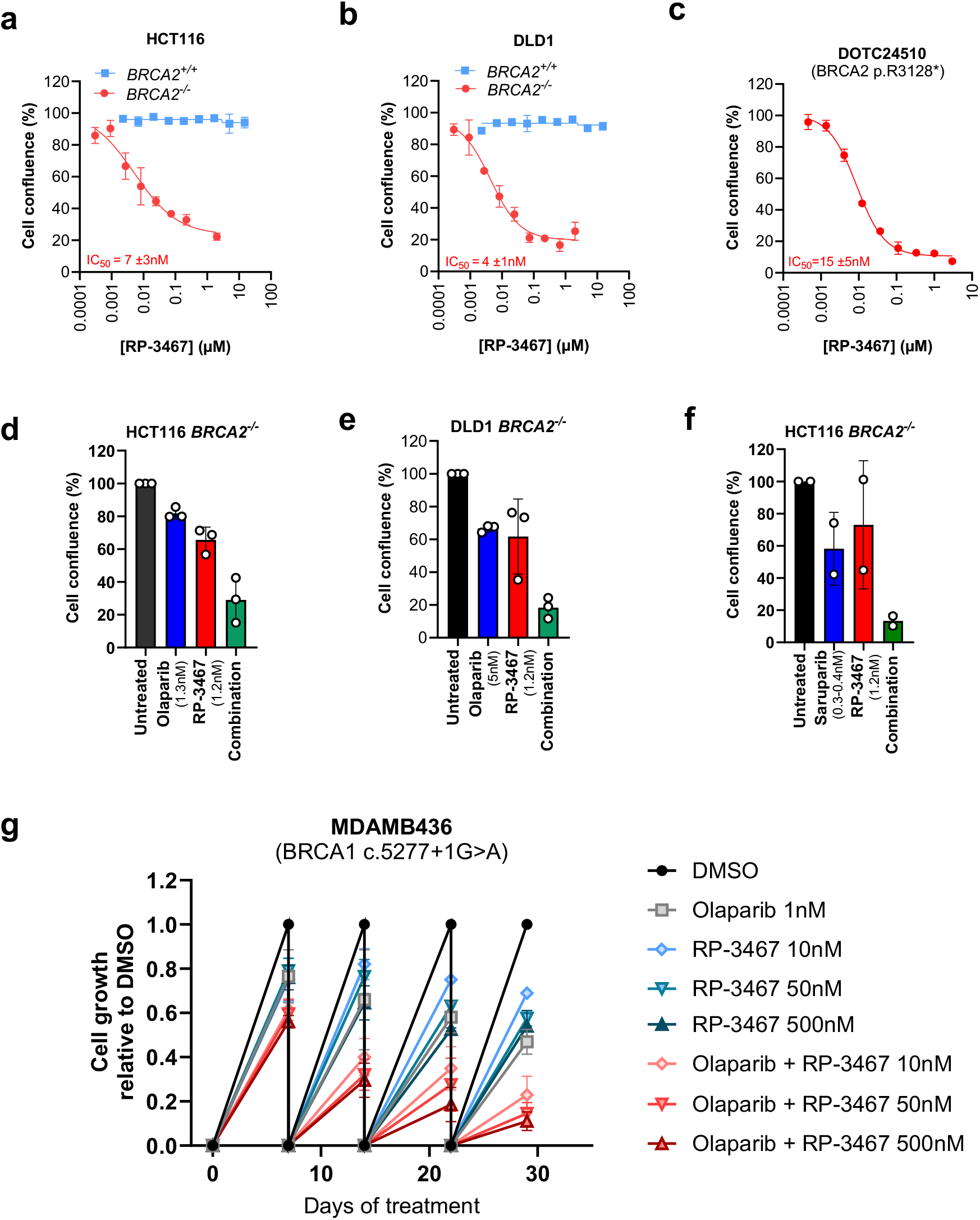
RP-3467 inhibits growth of HR-deficient cells as a single agent and elicits synergy with PARPi. **a-c.** Representative RP-3467 dose-response Incucyte cell growth assays in HCT116 *BRCA2^+/+^* and *BRCA2^-/-^* (a), DLD1 *BRCA2^+/+^* and *BRCA2^-/-^*(b) and DOTC24510 (*BRCA2*-mutant ovarian carcinoma; c) cells. *BRCA2^+/+^*cells: 5–7-day treatment. *BRCA2*-deficient cells: 10–14-day treatment. IC_50_ values are mean of 6,4, and 3 independent experiments, respectively, ±SD. **d-e.** Synergy between RP-3467 and olaparib in *BRCA2*-deficient cells. Cell growth (Incucyte) of HCT116 *BRCA2^-/-^* (d) and DLD1 *BRCA2^-/-^* (e) cells after indicated treatments. Symbols represent N=3 independent experiments with mean (bar) ±SD, normalized to DMSO. **f.** Synergy between RP-3467 and saruparib in HCT116 *BRCA2^-/-^*cells. Cell growth (Incucyte) after indicated treatments. Data from N=2 independent experiments (circles) with mean (bars) ±SD. **g.** Growth inhibition of MDAMB436 cells (*BRCA1-*mutant breast carcinoma) after 29-day continuous exposure to a combination of RP-3467 and the PARPi olaparib. Cells were grown in the presence of the indicated treatments and passaged every 7-8 days. At each time point, cells were reseeded at equal densities in all treatment arms. Cell numbers relative to DMSO at each subsequent collection time point are plotted. Data represent the mean of N=2 independent experiments ±SD.

We next tested whether RP-3467 enhances PARP inhibition in *BRCA2*-deficient cell lines. Co-treatment of HCT116 *BRCA2^-/-^,* DLD1 *BRCA2*^-/-^, or DOTC24510 cells with RP-3467 and the PARP1/2 inhibitor olaparib led to markedly reduced cell confluence compared to either agent alone, resulting in a synergistic combinatorial effect as measured by the ZIP synergy model^43^ (**Figure 2d,e, and Extended Data Figure 2a-d**). A similar combinatorial effect was observed with saruparib (AZD5305), a selective PARP1 inhibitor^44^, in HCT116 and DLD1 *BRCA2^-/-^* cells (**Figure 2f** and **Extended Data Figure 2e-g**). RP-3467 also showed synergy in HCT116 *BRCA2^-/-^* cells with two chemotherapeutic agents, carboplatin and SN-38 (active metabolite or the topoisomerase I poison irinotecan), as well as the antibody-drug conjugate payloads exatecan (topoisomerase I inhibitor) and SG3199 (DNA crosslinker; **Extended Data Figure 3**). Lastly, we assessed long-term treatment effects of RP-3467 in combination with olaparib in MDAMB436 cells, a *BRCA1*-mutant breast cancer line. Continuous exposure to RP-3467 in combination with olaparib resulted in sustained growth suppression over 29 days, particularly at higher concentrations of RP-3467, confirming durable synergy in a clinically relevant HRD background (**Figure 2g**).

### RP-3467 enhances the anti-tumor activity of PARP inhibitors in HRD xenograft and PDX models

To assess the anti-tumor potential of RP-3467 *in vivo*, we evaluated its efficacy alone and in combination with PARP inhibitors (PARPi) in multiple xenograft models of HRD cancer. In HCT116 *BRCA2^-/-^* xenografts, treatment with RP-3467 at 60 mg/kg or low-dose olaparib alone achieved modest tumor growth inhibition. In contrast, a combination of lower doses of RP-3467 with olaparib resulted in robust and durable tumor regression in a dose-dependent manner, without adversely affecting body weight (**Figure 3a, Extended Data Figure 4a**). The combination of 3 mg/kg RP-3467 with 25 mg/kg olaparib resulted in complete tumor clearance in 9 of 10 animals, with no evidence of regrowth following treatment cessation. Similarly, RP-3467 combination treatment was well tolerated and provided benefit even when olaparib was administered at its maximum tolerated dose of 100 mg/kg^45^, resulting in sustained tumor regressions and no combined effect on body weight (**Extended Data Figure 4b,c**).

**Figure 3.**
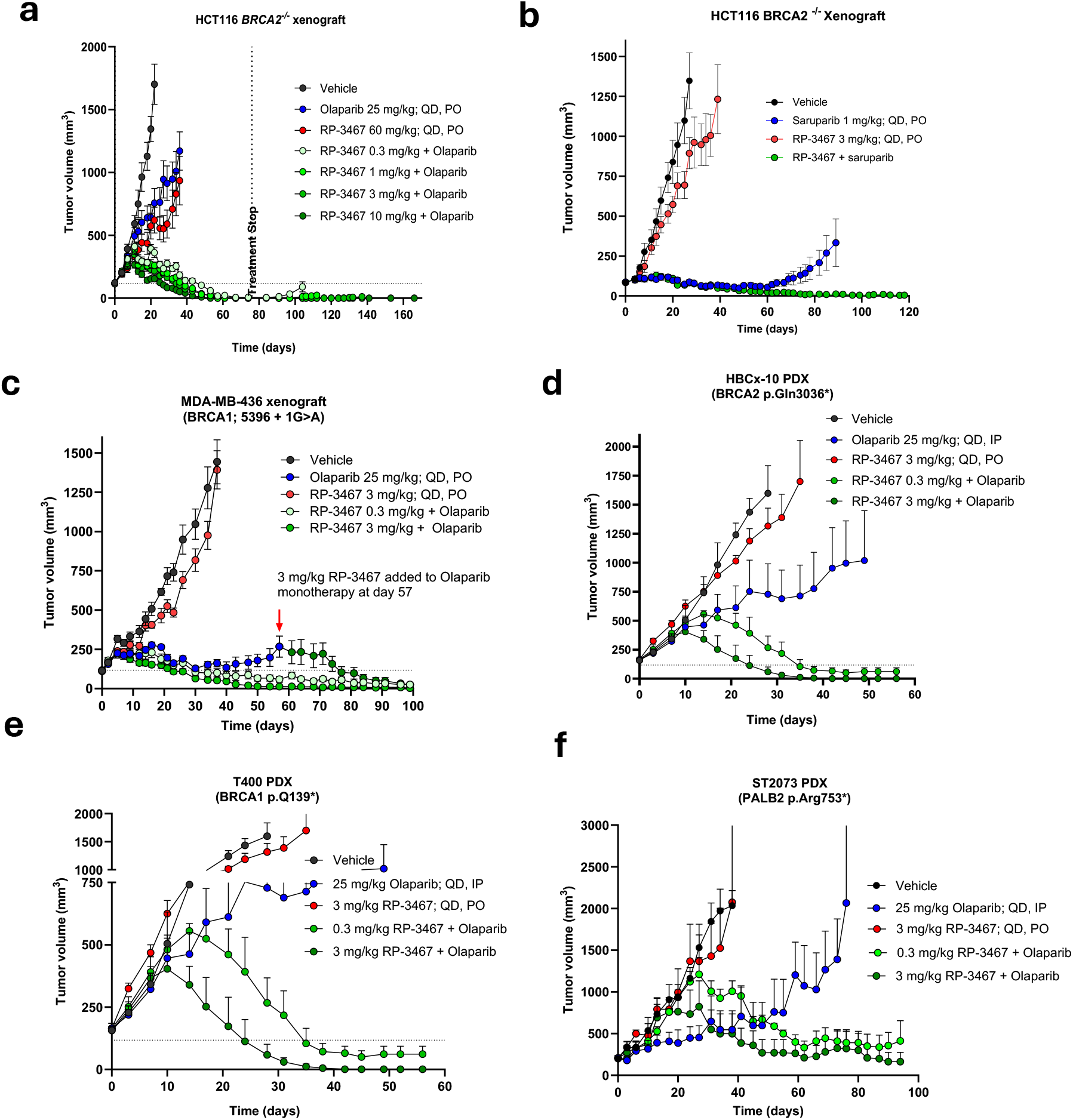
Combined antitumor activity of RP-3467 and PARP inhibitors in HRD tumor models *in vivo*. Tumor-bearing mice were treated with RP-3467, PARP inhibitors, or their combination to evaluate antitumor efficacy in HRD models. Treatment was initiated when tumors reached approximately 150–200 mm^3^. **a**. HCT116 *BRCA2^-/-^* xenografts were treated with varying doses of RP-3467 (0.3-10 mg/kg) alone or in combination with olaparib (25 mg/kg). *N*=9-10 mice/group. **b**, HCT116 *BRCA2^-/-^* xenografts were treated with RP-3467 (3 mg/kg) and/or the PARP1-inhibitor saruparib (0.03–0.1 mg/kg, QD) to assess combinatorial activity. *N*=7-8 mice/group. **c**, MDA-MB-436 (*BRCA1^-/-^*) xenografts were treated with RP-3467 (3 mg/kg), olaparib (25 mg/kg), or both. In one cohort, RP-3467 was added at day 57 to mice that had previously received olaparib monotherapy. *N*=6-10 mice/group. **d**. Patient-derived xenograft model HBCx-22 was treated with RP-3467 (0.3 or 3 mg/kg), olaparib (50 mg/kg QD), or the combination. *N*=3 mice/group. **e**. T400 PDX model was treated with RP-3467 (3 mg/kg), olaparib (25 mg/kg), or both, administered daily. *N*=3 mice/group. **f**. ST2073 PDX model was treated with RP-3467 (0.3 or 3 mg/kg), olaparib (25 mg/kg, QD or IP), or their combination. Data reported as mean tumor volume ±SEM for each group. *N*=3 mice/group.

RP-3467 also showed enhanced combinatorial activity with the PARP1-selective inhibitor saruparib in HCT116 *BRCA2^-/-^* xenografts, both at the reported maximally efficacious dose^44^ (1 mg/kg; **Figure 3b**) as well as at lower doses (**Extended Data Fig. 4d**). The combination with saruparib had no additional effect on mouse body weight (**Extended Data Figure 4e**). As expected, RP-3467 showed no activity as monotherapy or in combination in *BRCA2*-proficient HCT116 xenografts, confirming its selectivity for HRD tumors (**Extended Data Figure 4f**). To extend these findings to a *BRCA1*-deficient context, we assessed the RP-3467/olaparib combination in MDA-MB-436 xenografts (*BRCA1* c.5396+1G>A). As in the *BRCA2*-deficient model, combination therapy resulted in pronounced tumor regression (**Figure 3c**). In tumors that initially responded and then progressed on olaparib monotherapy, the addition of RP-3467 was sufficient to restore drug sensitivity and re-induce regressions, suggesting that Polθ inhibition can potentially prevent and/or overcome acquired PARPi resistance.

We extended these findings by examining RP-3467-olaparib combinations in HRD patient-derived xenografts (PDX). RP-3467 combined with olaparib resulted in greater antitumor efficacy than olaparib alone in HBC-x10 TNBC (triple negative breast cancer cell line with *BRCA2* p.Gln3036*) (**Figure 3d**), T-400 TNBC (*BRCA1* p.Q139*) (**Figure 3e**), and ST2073 uterine carcinoma (*PALB2* p.Arg753*) PDX (**Figure 3f**). These findings underscore the synergistic antitumor activity of Polθ ATPase inhibition when combined with PARP inhibitors across multiple HR-deficient tumor models.

Beyond PARPi, RP-3467 also potentiated the activity of several DNA-damaging agents, including irinotecan and carboplatin, in both HCT116 and DLD1 *BRCA2^-/-^* models (**Extended Data Figure 5a-d**), suggesting that Polθ inhibition may enhance the broader sensitivity of HRD tumors to genotoxic chemotherapy. These combinations were similarly well tolerated, with no significant body weight loss over the course of the studies (**Extended Data Figure 5a-d**). Lastly, in OVCAR3 xenografts, a *BRCA1/2* wild-type ovarian cancer model, RP-3467 in combination with Dato-DXd (a TROP2-directed ADC) produced additive tumor control (**Extended Data Figure 5e**).

Collectively, these data establish RP-3467 as a potent Polθ ATPase inhibitor with robust *in vivo* activity against HRD tumors in rational combination regimens. Notably, combining Polθ and PARP inhibitors leads to deep and durable tumor regressions, supporting a therapeutic strategy that targets complementary arms of the DNA repair network to maximize synthetic lethality and overcome resistance.

### RP-3467 does not exacerbate hematological toxicity associated with PARPi treatment

Dosing of PARPi in the clinic is limited by hematologic toxicities, particularly anemia, thrombocytopenia, and neutropenia. To assess whether RP-3467 impacts hematologic tolerability, we evaluated blood cell counts in non-tumor-bearing CD-1 mice treated for 5 weeks. Hematologic profiling revealed that RP-3467, whether administered alone or in combination with olaparib, had minimal impact on red blood cell (RBC) counts, lymphocytes, neutrophils, platelets, eosinophils, and reticulocytes (**Figure 4a-f**). As expected and consistent with its known myelosuppressive effects, olaparib monotherapy significantly affected several hematological parameters - most notably platelet, RBC, and neutrophil counts. However, these effects were not worsened by co-administration with RP-3467. These results indicate that RP-3467 does not compromise bone marrow function in mice at therapeutically active doses and, significantly, does not exacerbate hematologic toxicity associated with PARPi, supporting the potential of co-targeting Polθ and PARP1/2 in clinical settings.

**Figure 4.**
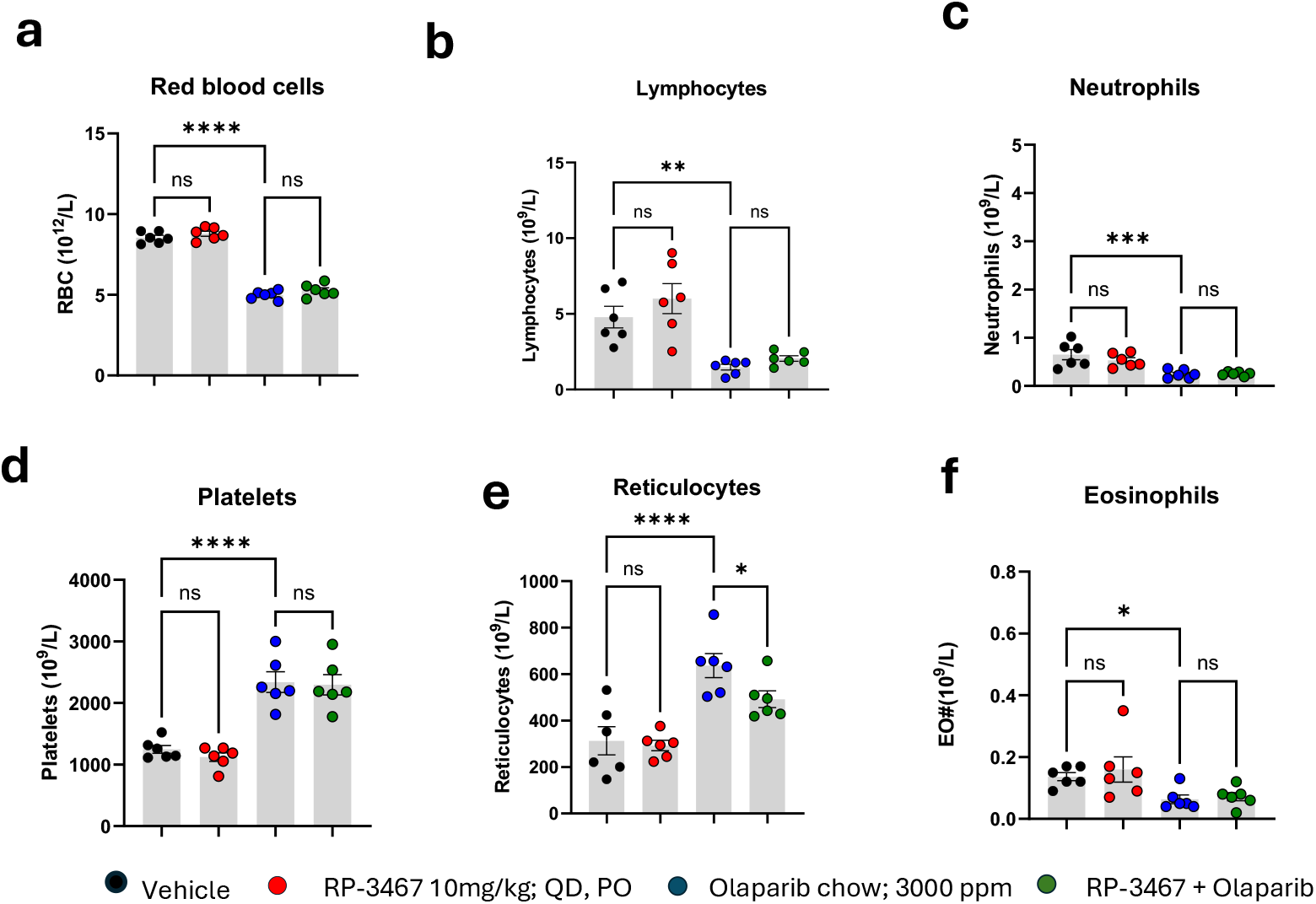
Effects of RP-3467 and olaparib on blood cell parameters. Tumor-naïve CD-1 female mice were treated daily PO with RP-3467, olaparib-formulated chow, or the combination for 5 weeks. **a-f**. Hemoglobin (a) and complete blood counts (b-f) are expressed as mean +/- SEM (bars) from N=5-6 mice per group (symbols). The color legend is shown below the figure panels. Dotted lines represent values for CRL female CD-1 mice of similar age. One-way ANOVA evaluated statistical differences with Fisher’s LSD test. Ns = not significant; * P < 0.05, ** P < 0.001; **** P < 0.0001.

### RP-3467 treatment disrupts mitotic DNA repair *in vivo*

To corroborate that the efficacy of RP-3467 in mouse xenografts is driven by Polθ inhibition, we assessed the pharmacodynamic effects of RP-3467 *in vivo* and analyzed biomarkers of mitotic DNA damage in HCT116 *BRCA2^-/-^* xenografts. Immunofluorescence analysis of formalin-fixed and paraffin-embedded (FFPE) tumor sections revealed a significant increase in micronucleated cells upon treatment with either RP-3467 or olaparib monotherapy, with an additive increase observed in combination (**Figure 5a,b**), suggesting enhanced genomic instability. We next examined mitotic cells for accumulation of CIP2A foci to visualize DNA lesions in mitosis. Tumors were stained for phospho-histone H3 (H3pS10) to mark mitotic cells and for CIP2A to detect DNA damage-associated foci. While RP-3467 and olaparib each increased the frequency of CIP2A-positive mitoses, the combination induced the highest percentage of cells with more than 10 discrete CIP2A foci (**Figure 5c,d**). These findings show that RP-3467 increases the persistence of DNA lesions during mitosis, consistent with its role in disrupting the mitotic MMEJ pathway, which typically resolves replication-associated breaks in HR-deficient cells.

**Figure 5.**
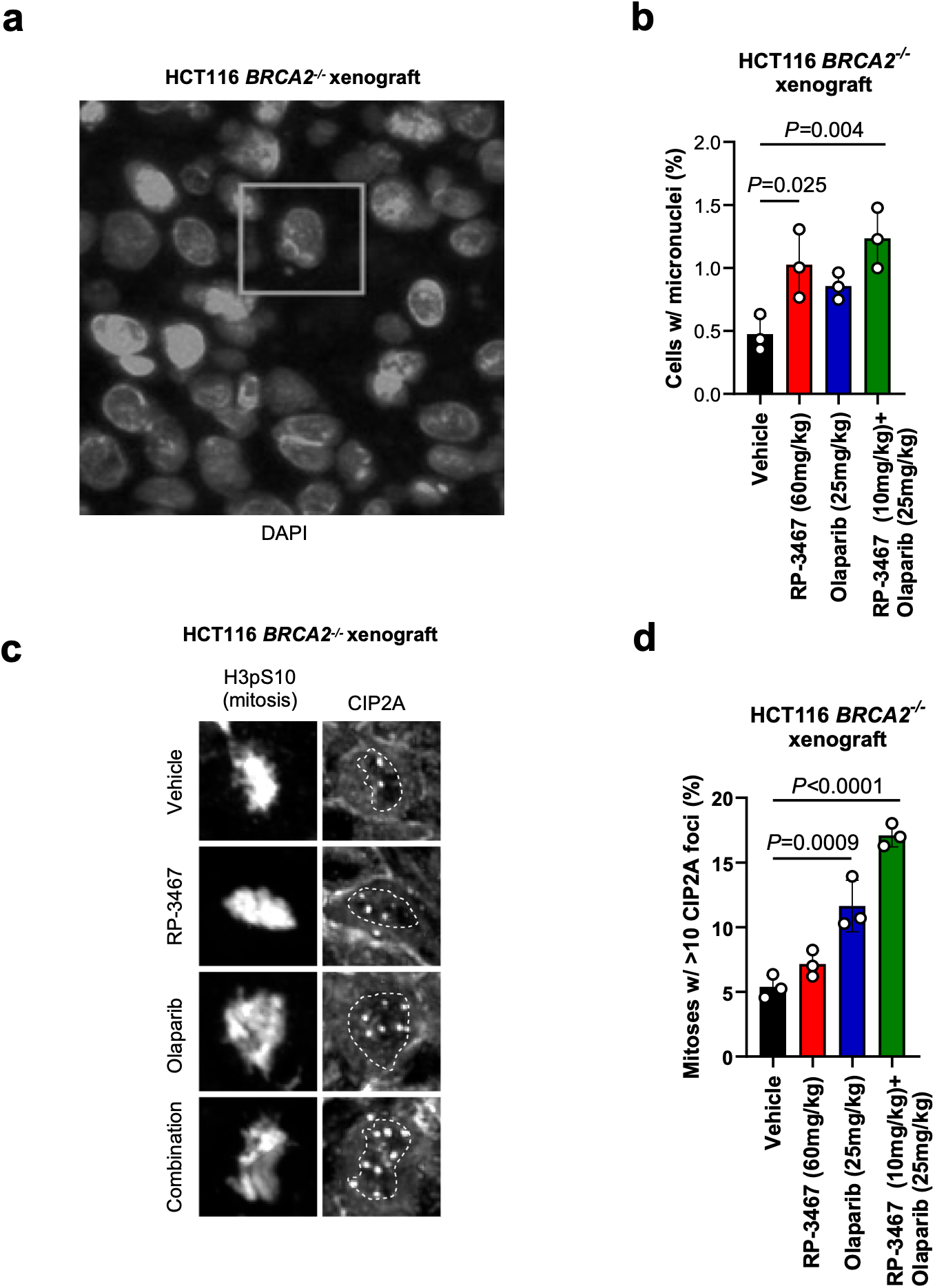
Pharmacodynamics of RP-3467 *in vivo*. **a**. Example image of DAPI-stained nuclei in an HCT116 *BRCA2^-/-^* xenograft tumor. A micronucleus-containing cell is highlighted. **b.** Quantification of micronuclei-containing cells in HCT116 *BRCA2^-/-^* tumors after indicated treatments. **c.** Example images of mitotic cells in HCT116 *BRCA2^-/-^* xenograft tumors processed for immunofluorescence with anti-H3pS10 (mitotic marker) and anti-CIP2A antibodies after indicated treatments. **d.** Quantification of mitotic cells containing >10 CIP2A foci in HCT116 *BRCA2^-/-^* tumors upon indicated treatments. Data in **b,d** are N=3 tumors/group (circles) with mean (bars) ±SD. P-values calculated with one-way ANOVA.

### Clinical evidence of efficacy in a patient with a *BRCA1*-mutant ovarian cancer and an acquired *TP53BP1* loss-of-function mutation

The clinical potential of Polθ inhibition was evaluated in a first-in-human Phase I POLAR trial (NCT06560632), which assessed the safety, tolerability, and preliminary antitumor activity of RP-3467 as monotherapy or in combination with the PARP inhibitor, olaparib. Among the first patients enrolled in the combination arm was a woman in her 70’s with metastatic high-grade serous ovarian cancer harboring a pathogenic germline *BRCA1* mutation (c.798_799delTT, p.Ser267Lysfs*19). The patient initially received carboplatin, paclitaxel, and bevacizumab, which produced a clinical response. The patient subsequently underwent primary debulking surgery, and olaparib treatment was then initiated as maintenance therapy. Despite this treatment, she developed oligoprogression approximately four months after completing her platinum regimen, at which point she had become platinum-resistant. This progression resulted in secondary debulking surgery, after which olaparib was resumed postoperatively. Following radiologic progression on olaparib, the patient was sequentially treated with pegylated doxorubicin, paclitaxel, and DB-1305 (a TROP-2-directed antibody-drug conjugate) (**Figure 6a**).

**Figure 6.**
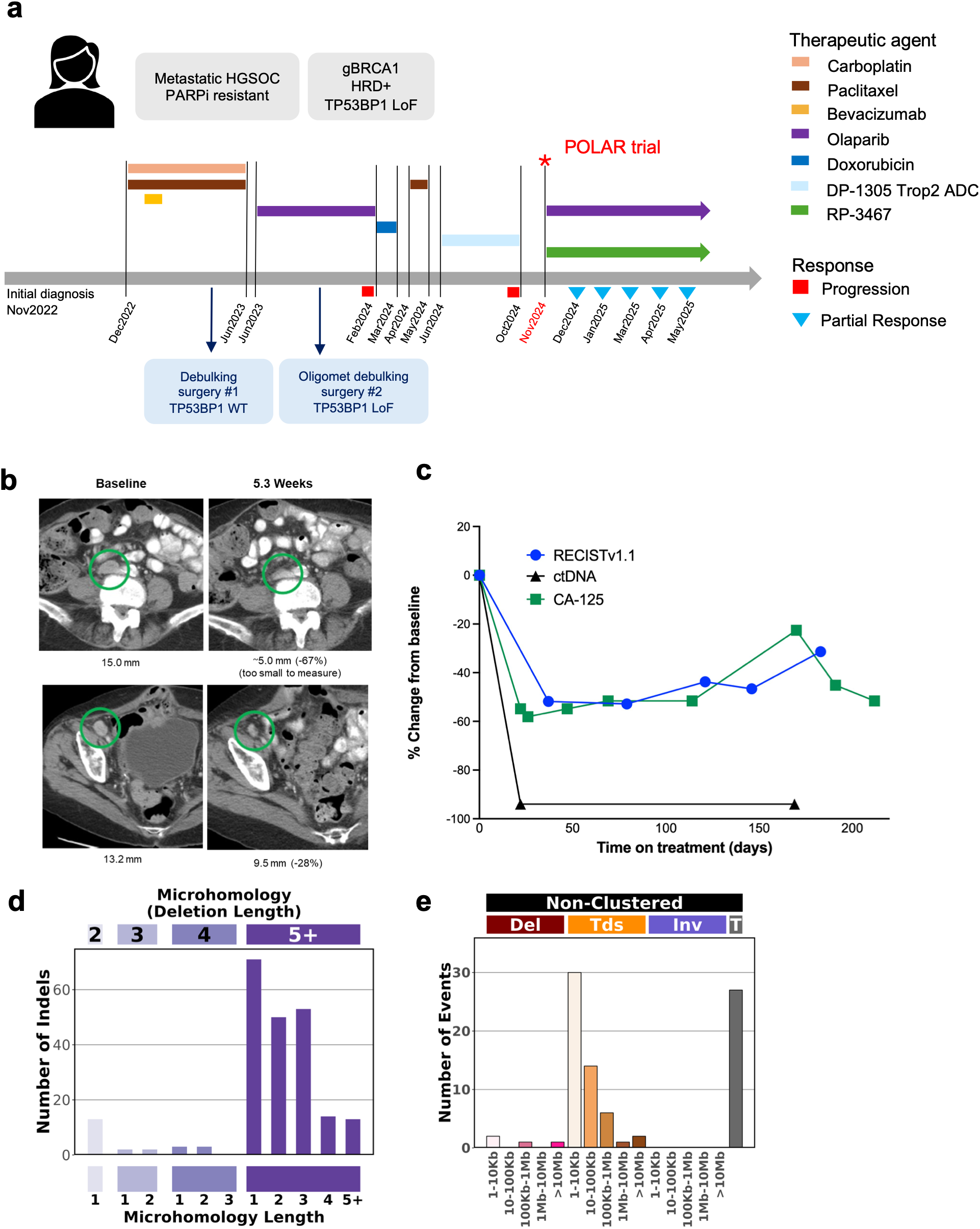
Clinical response to combined PARP and Polθ inhibition in a germline *BRCA1*-mutant ovarian cancer patient. **a.** Treatment timeline showing therapy history from diagnosis through the POLAR trial. A female in her 70’s with metastatic HGSOC (g*BRCA1*, HRD+, *TP53BP1* LoF) received multiple treatment lines (colored bars). Red circles indicate progression; blue triangles indicate partial response. Two debulking surgeries identified a *TP53BP1* wild-type tumor (surgery #1) and a *TP53BP1* P1846fs mutated tumor (surgery #2). **b.** Serial CT scans of a representative lymph node lesion (green circles) at baseline and 5.3 weeks post-RP-3467 treatment, showing tumor reduction from 15.9 mm to 6.1 mm (62%, below RECIST threshold), with subsequent measurements of 13.2 mm and 9.5 mm (28% reduction). **c.** Longitudinal biomarker changes during treatment. RECIST v1.1 measurements (blue), ctDNA (black), and CA-125 (green) plotted as percent change from baseline. ctDNA remained undetectable (-100%) throughout treatment, while RECIST and CA-125 showed initial response followed by gradual increases. **d.** Surgical timeline illustrating clonal evolution from *TP53BP1* wild type (surgery #1, pink) to *TP53BP1* p.1846fs mutant tumor (surgery #2, blue). **e.** Deletion length distribution grouped by microhomology length at junctions (2-5+ bp). Enrichment of 3-4 bp microhomology indicates MMEJ pathway activity downstream of the germline *BRCA1* mutation. **f.** Distribution of non-clustered structural variants by type (deletions, tandem duplications, inversions) and microhomology length. Enrichment of deletions with longer microhomology signatures supports active Polθ-mediated MMEJ repair.

The patient then enrolled in the POLAR trial and initiated treatment with RP-3467 (80 mg QD) combined with olaparib (200 mg BID). No dose-limiting toxicities were observed during the first cycle. However, treatment-emergent myelosuppression necessitated a dose reduction of RP-3467 to 40 mg daily. This regimen was well tolerated, and the patient achieved a confirmed partial response (PR; RECISTv1.1; sum of target lesions -51.8% from baseline) after 37 days of treatment, which was sustained for 17 weeks (**Figure 6b,c**). Although the patient experienced radiographic disease progression at 26.1 weeks, she was treated beyond progression due to continued clinical benefit and remains on therapy at 38.1 weeks. Notably, CA-125 levels mirrored the radiographic response, remaining within the normal range throughout treatment. Circulating tumor DNA (ctDNA) levels showed a near-complete reduction (94%) by day 22, preceding the radiographic response, and remained suppressed through week 24 (**Figure 6b,c**).

Baseline pre-dose ctDNA analyses confirmed the g*BRCA1* p.Ser267Lysfs*19 variant. In addition, we identified a loss-of-function mutation in *TP53BP1* (p1846fs), a well-established driver of PARP inhibitor resistance and a known sensitizer to Polθ inhibition that has been extensively characterized in preclinical models and detected in a limited number of clinical cases^30,46^. To determine when this *TP53BP1* mutation emerged relative to PARPi treatment, whole-genome sequencing (WGS) was performed on two surgical samples: one collected before PARPi therapy and the other at the time of oligoprogression on olaparib (**Figure 6d**). As expected for a g*BRCA1* tumor, both samples showed HRD signatures (**Figure 6e-f**). However, the *TP53BP1* mutation was detected only in the sample acquired post-PARPi treatment (**Figure 6d and Extended data Figure 6**), suggesting that it likely emerged as a resistance mechanism to PARP inhibitor therapy. WGS analysis of the patient’s tumor samples identified a pattern of frequent deletions in areas with microhomology and tandem duplications of 1-100Kb, indicative of HRD **(Figure 6e-f and Extended data Figure 7a-e**).

In conclusion, this case study illustrates the clinical potential of RP-3467 to overcome PARP inhibitor resistance in HRD tumors, particularly those harboring *TP53BP1* loss. More broadly, they underscore the therapeutic value of targeting Polθ inhibition to expand treatment options for a growing patient population.

## DISCUSSION

The clinical success of PARP inhibitors in HRD tumors has established synthetic lethality as a powerful therapeutic paradigm. However, the benefit of PARPi is often limited by intrinsic and acquired resistance, as well as hematological toxicities that constrain combination strategies^13,14^. Our study identifies RP-3467 as a selective, potent, and well-tolerated inhibitor of the helicase-like ATPase domain of polymerase theta (Polθ), an essential enzyme in the MMEJ pathway, and demonstrates its ability to enhance and prolong PARPi efficacy in HR-deficient tumors.

Mechanistically, RP-3467 disrupts Polθ’s ATPase activity, leading to MMEJ suppression and accumulation of unresolved mitotic DNA damage in HRD cells. Consistent with the established role of Polθ in mitotic repair^24^, RP-3467 treatment increased micronuclei formation and CIP2A-marked foci in mitosis, suggestive of impaired resolution of replication-associated DNA lesions. These effects were observed at nanomolar concentrations that correlated with cellular target engagement, confirming that Polθ helicase activity is indispensable for maintaining genome stability under conditions of HR loss. Pharmacologic inhibition of Polθ with RP-3467 induced synthetic lethality in multiple *BRCA1/2*-deficient models and exhibited synergy with PARP1/2-(olaparib) inhibitors, as well as multiple chemotherapy agents. Notably, co-treatment with RP-3467 and PARPi resulted in deep and durable tumor regressions across cell line- and patient-derived xenograft models from breast, ovarian, and other cancers carrying *BRCA1/2* or *PALB2* mutations, without evidence of exacerbated systemic or hematologic toxicity compared to PARPi monotherapy. These findings underscore the therapeutic potential of co-targeting Polθ and PARP1/2 as a strategy to amplify synthetic lethality and overcome limitations of currently tested combination therapies.

Our work builds on prior studies demonstrating the vulnerability of HRD tumors to Polθ inhibition^27–29,32^ and extends this concept to the helicase-like domain, which has emerged as a mechanistically distinct and druggable node within the MMEJ machinery. Unlike polymerase-domain inhibitors such as ART558^32^, which block Polθ’s gap-filling DNA synthesis, RP-3467 targets the helicase-like ATPase domain and interferes with the upstream processing of resected DNA ends required to initiate MMEJ. Furthermore, in contrast to novobiocin, a repurposed antibiotic with broader chaperone and ATPase activity^35^, RP-3467 is a highly selective Polθ ATPase inhibitor optimized for potency, target engagement, and tolerability, enabling sustained combination with PARP inhibitors. Whereas PARP inhibitor combinations with other DDR agents frequently lead to exacerbated hematologic toxicity^13,19,48^, RP-3467 did not increase the myelosuppressive effects of PARPi in mouse studies, supporting its suitability for sustained combination therapy. Clinically, the patient treated with RP-3467 (40 mg QD) combined with olaparib (200 mg BID) showed no treatment-related hematologic toxicity, consistent with the favorable preclinical safety profile. Our study provides initial clinical evidence for Polθ helicase inhibition as a strategy to re-sensitize some tumors with acquired PARPi resistance. Specifically, a patient with *BRCA1*-mutant ovarian cancer harboring a *TP53BP1* mutation, which is a known PARPi resistance mechanism^14,46^, RP-3467 combined with olaparib induced both a radiographic and molecular response. This observation supports preclinical findings that *BRCA1/TP53BP1* double-deficient tumors retain dependency on MMEJ and remain vulnerable to Polθ inhibition^30,32^, thereby validating RP-3467 as a potential intervention in specific post-PARPi settings.

Reversion mutations restoring *BRCA1/2* function are a common mechanism of PARPi resistance in human tumors and often display features of MMEJ, such as microhomology at breakpoint junctions^16–18,49,50^. These signatures have led to the suggestion that MMEJ activity underlies the acquisition of reversion mutations and resistance to PARPi^49^. In this context, inhibition of Polθ may not only potentiate PARPi efficacy but also delay or prevent the emergence of reversion-based resistance by suppressing the mutagenic repair processes that facilitate it. Thus, combination therapy with RP-3467 and PARPi may offer a dual benefit: enhanced depth of response and improved durability by impeding the evolution of resistance at its source.

Together, these data establish RP-3467 as a potent and selective inhibitor of Polθ ATPase activity and provide a compelling rationale for clinical development in HRD tumors. Beyond synthetic lethality, the ability of RP-3467 to restore sensitivity to a patient who previously progressed on PARPi suggests broader utility in overcoming acquired resistance mechanisms, particularly those involving loss of 53BP1/Shieldin. As the field seeks to extend the benefits of PARP-targeted therapy, Polθ helicase inhibition emerges as a promising, clinically actionable strategy to expand the therapeutic window and the durability of response in DNA repair-deficient cancers.

## ACKNOWLEDGEMENTS

We thank Inocras Inc. for whole-genome sequencing, processing, and analytical support for our clinical samples. Polθ-related work in the Sfeir lab is supported by grants from the NIH/NCI (R01CA229161, R01CA294696, P50CA247749) and the The V Foundation for Cancer Research. T.A.Y. holds the Ransom Horne, Jr. Endowed Professorship for Cancer Research at The University of Texas MD Anderson Cancer Center and is supported by National Cancer Center Support Grant CA016672, awarded to The University of Texas MD Anderson Cancer Center; US Department of Defense grants W81XWH2210504_BC211174 and W81XWH-21-1-0282<u>_</u>OC200482; V Foundation Scholar Grant VC2020-001; National Institutes of Health R01 1R01CA255074 grant and The University of Texas MD Anderson Cancer Center National Institutes of Health SPORE in Ovarian Cancer 1P50CA281701-01.

## ETHICS STATEMENT

This study was reviewed and approved by the Memorial Sloan Kettering Cancer Center Institutional Review Board/Privacy Board (Memorial Sloan Kettering Cancer Center, New York, NY, USA) under protocol number 24-316, with approval granted on October 15, 2024. All participants provided written informed consent prior to enrollment. The study was conducted in accordance with the Declaration of Helsinki.

## AUTHOR CONTRIBUTION

A.S., S.J.M., E.Y.R., M. Zimmermann: Conceptualization, experimental data collection and analysis, paper writing. M.-C.M., G.F., P.M., C.G., H.P., S.F., D.H., H.K., R.P., S.Y.Y., M.-E.L., R.H., P.N., J.D.S., K.E.Z., I.K., D.U., A.V., R.G., N.M., P.B.: Experimental data collection and/or analysis. W.C.B., M.G., V.R., T.A.Y., M.K., M. Zinda, A.R.: Supervision and conceptualization.

## EXTENDED DATA FIGURE LEGENDS

**Extended Data Figure 1.**
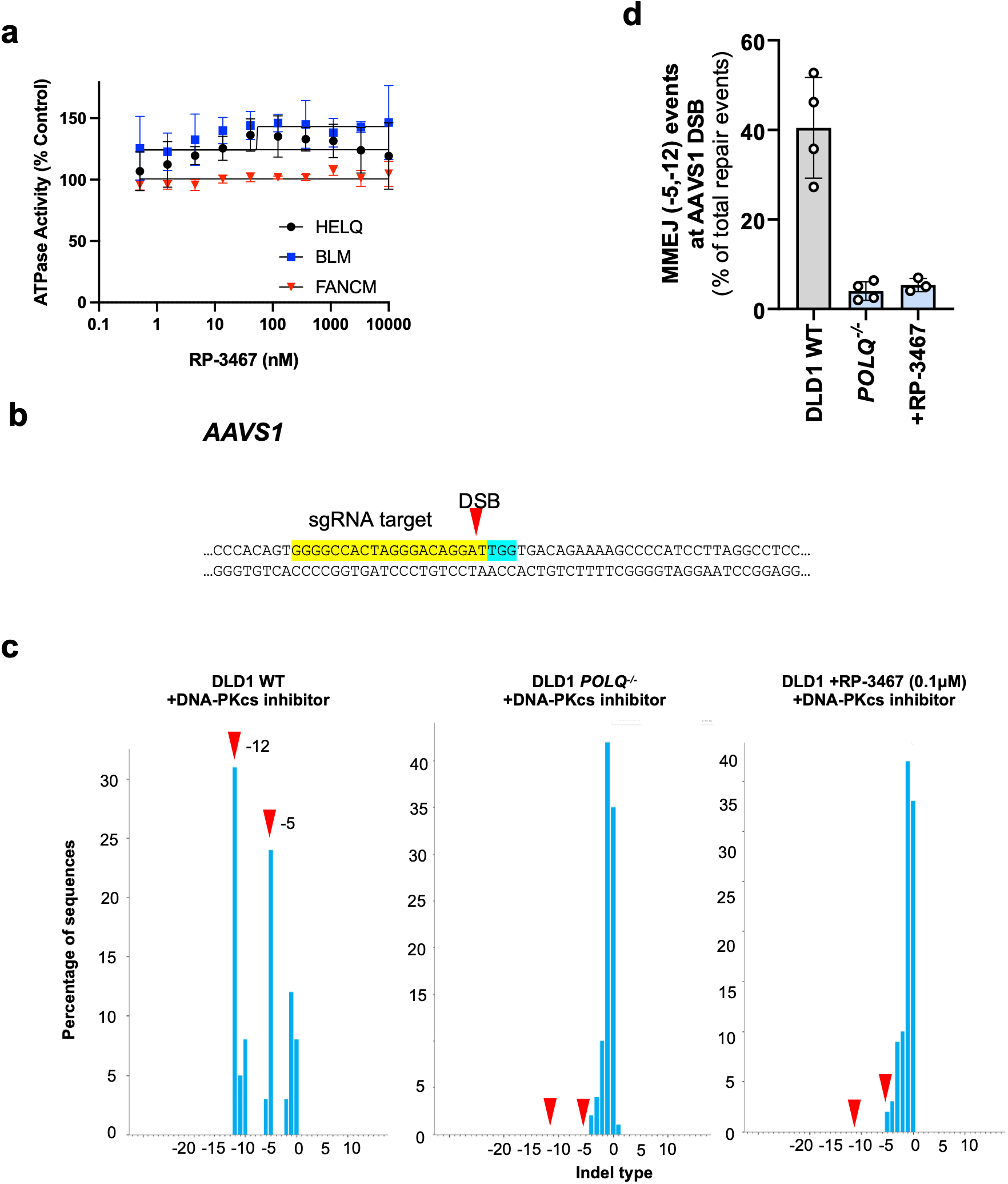
Related to Figure 1. **a. RP-3467 does not inhibit other DNA-dependent ATPases**. ADPglo dose-response curves measuring inhibition of indicated recombinant ATPases by RP-3467. **b-d. Validation of MMEJ DSB repair assay. b.** Sequence of a targeted double-strand break (DSB) site in the AAVS1 locus. sgRNA target and PAM are highlighted. **c.** Example indel quantification plots from DLD1 WT, DLD1 *POLQ^-/-,^* and RP-3467-treated DLD1 WT cells (all in the presence of a DNA-PKcs inhibitor). *AAVS1* DSB was induced by Cas9/sgRNA, the breakpoint site was PCR-amplified and Sanger-sequenced. Sequencing traces were deconvolved with ICE (https://ice.editco.bio/). Polθ-dependent -5 and -12 indels are highlighted. **d**. Quantification of -5 and -12 (MMEJ) indel events in DLD1 WT, DLD1 *POLQ^-/-^,* and RP-3467-treated DLD1 WT cells (in the presence of DNA-PKcs inhibitor). Data from *N*=3-4 independent experiments (circles) with mean (bars) ±SD.

**Extended Data Figure 2:**
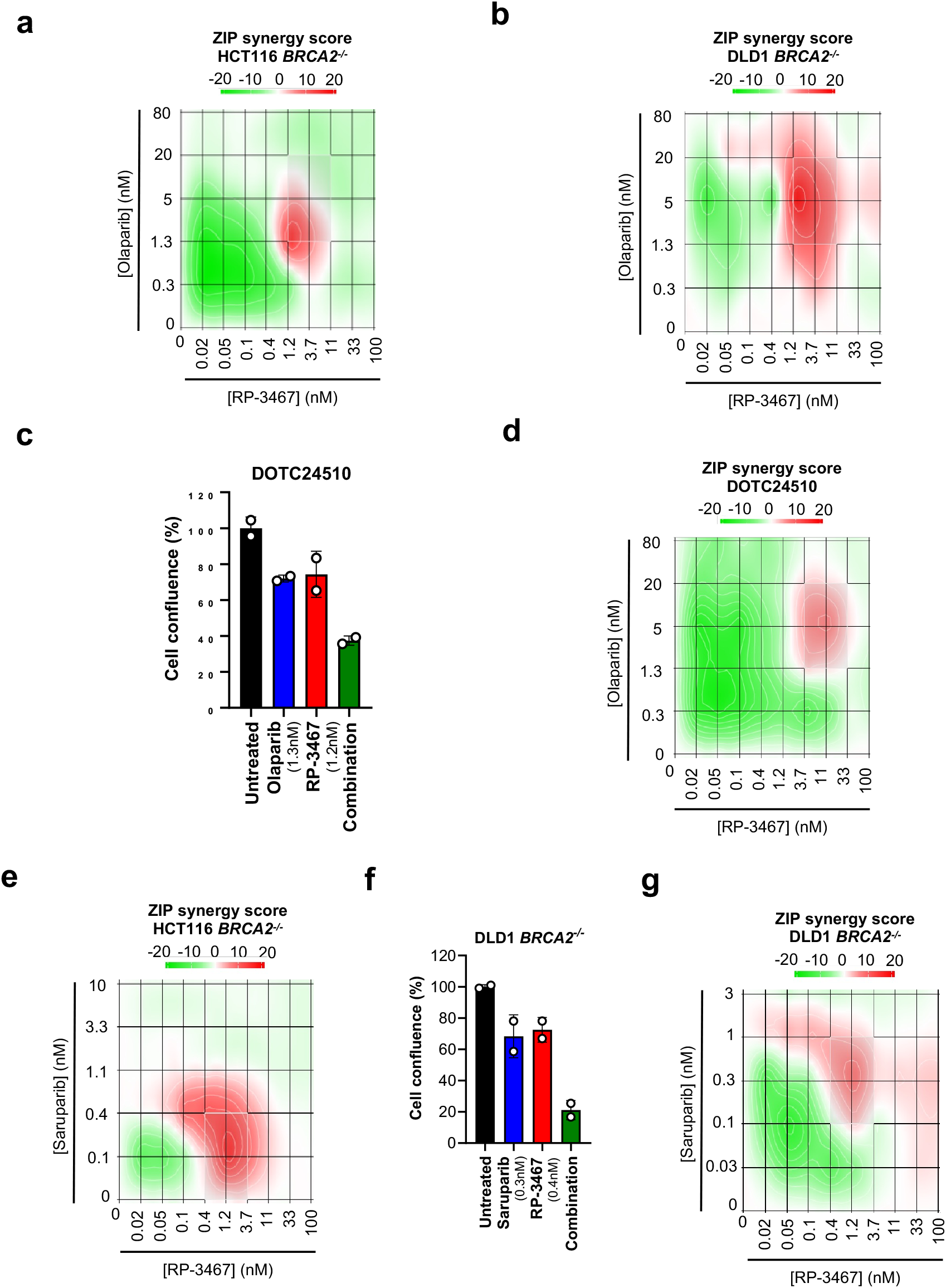
Synergy between RP-3467 and PARPi in HR-deficient cell lines. Related to Figure 2. **a,b.** Representative (of N=3 independent experiments) ZIP synergy plots at indicated concentrations of RP-3467 and olaparib in HCT116 *BRCA2^-/-^* (a), DLD1 *BRCA2^-/-^* (b) cells. **c.** Cell growth (Incucyte) of DOTC24510 (*BRCA2-*mutant ovarian carcinoma) cells after indicated treatments. Symbols represent N=2 technical replicates with mean (bar) ±SD, normalized to DMSO. **d.** ZIP synergy plot at indicated concentrations of RP-3467 and olaparib in DOTC24510 cells. **e.** Representative (of N=2 independent experiments) ZIP synergy plot at indicated concentrations of RP-3467 and saruparib in HCT116 *BRCA2-/-* cells. **f.** Cell growth (Incucyte) of DLD1 *BRCA2^-/-^* cells after indicated treatments. Symbols represent N=2 technical replicates with mean (bar) ±SD, normalized to DMSO. **g.** ZIP synergy plot at indicated concentrations of RP-3467 and saruparib in DLD1 *BRCA2^-/-^* cells. In a,b,d,e,g, score >10 (red color) indicates synergy.

**Extended Data Figure 3.**
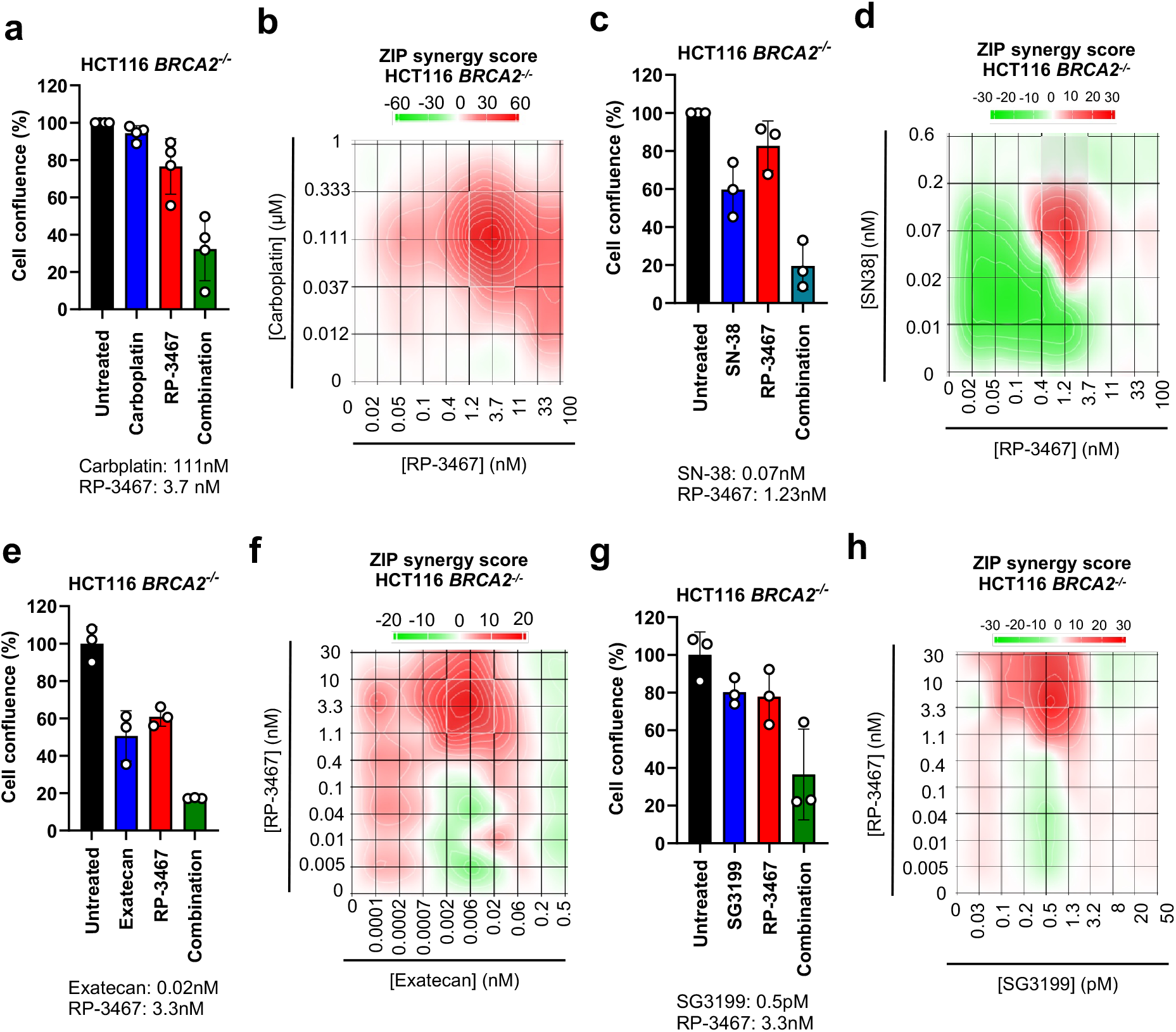
Synergy between RP-3467 and chemotherapeutic agents in HR-deficient cell lines. Related to Figure 2. **a,c,e,h.** Cell growth (Incucyte) of HCT116 *BRCA2^-/-^* cells after indicated treatments. Data from N=3-4 independent experiments (circles) with mean (bars) ±SD. **b,d,f,h.** Representative ZIP synergy plots at indicated compound concentrations in HCT116 *BRCA2^-/-^* cells. Score >10 (red color) indicates synergy.

**Extended data figure 4.**
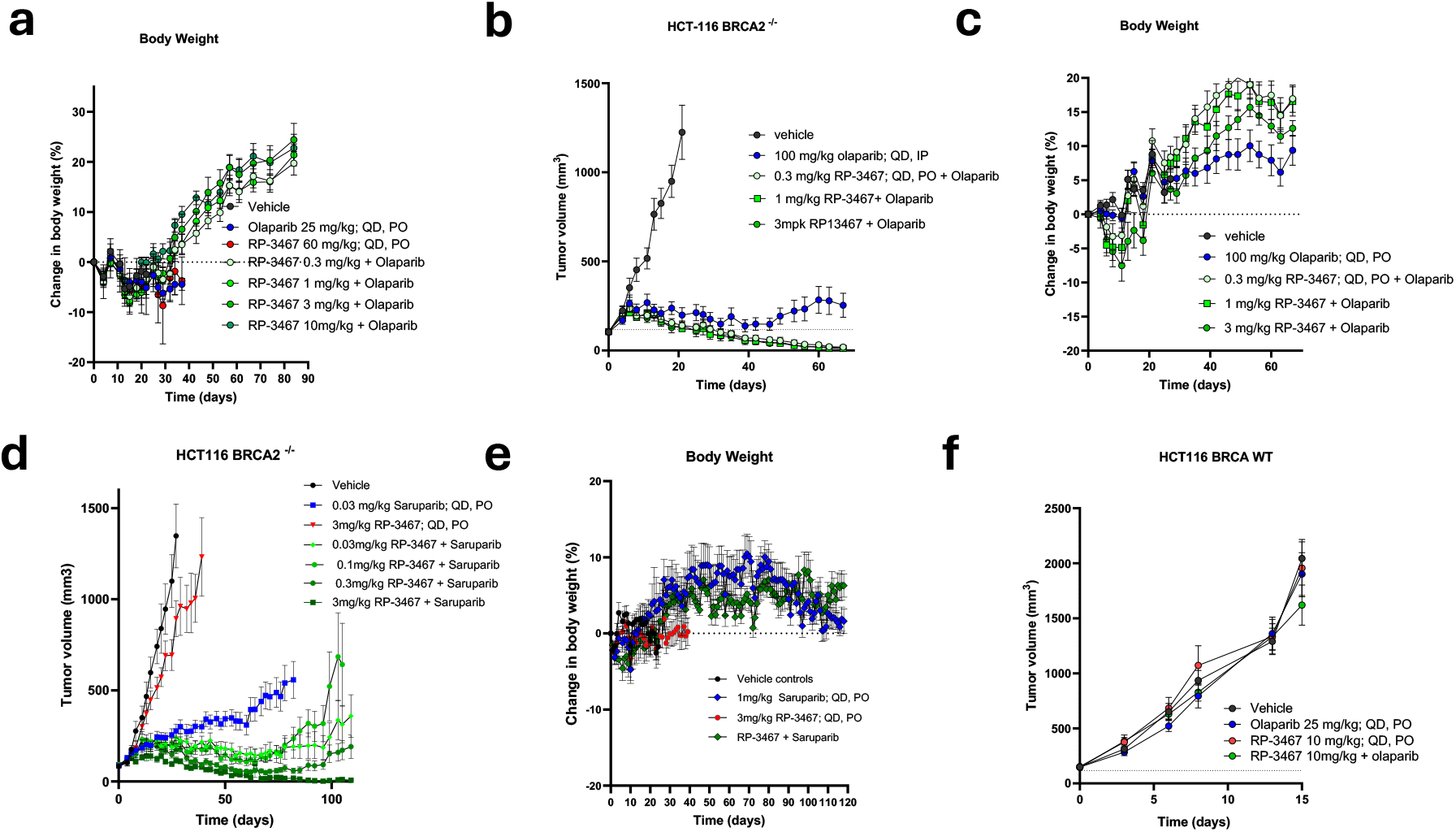
*In vivo* efficacy and tolerability of RP-3467 in tumor-bearing mice. Related to Figure 3. **a.** Body weights of mice treated with low dose olaparib and RP-3467. **b,c.** Tumor xenograft volume (**b**) and mouse body weights (**c**) of HCT116 *BRCA2^-/-^* tumor-bearing mice treated with RP-3467 in combination with 100 mg/kg olaparib. **d.** Tumor xenograft volume from HCT116 *BRCA2^-/-^* tumor-bearing mice treated with RP-3467 in combination with 0.03 mg/kg saruparib. **e.** Body weights of HCT116 *BRCA2^-/-^* tumor-bearing mice treated with RP-3467 and 1 mg/kg saruparib. **f.** Tumor xenograft volume from HCT116 *BRCA2^+/+^* tumor-bearing mice treated with RP-3467 in combination with olaparib. Data in all panels are mean ± SEM; *N*=5-10 mice/group. Mice were treated orally with RP-3467, olaparib or saruparib once daily for the duration of the experiments.

**Extended Data Figure 5.**
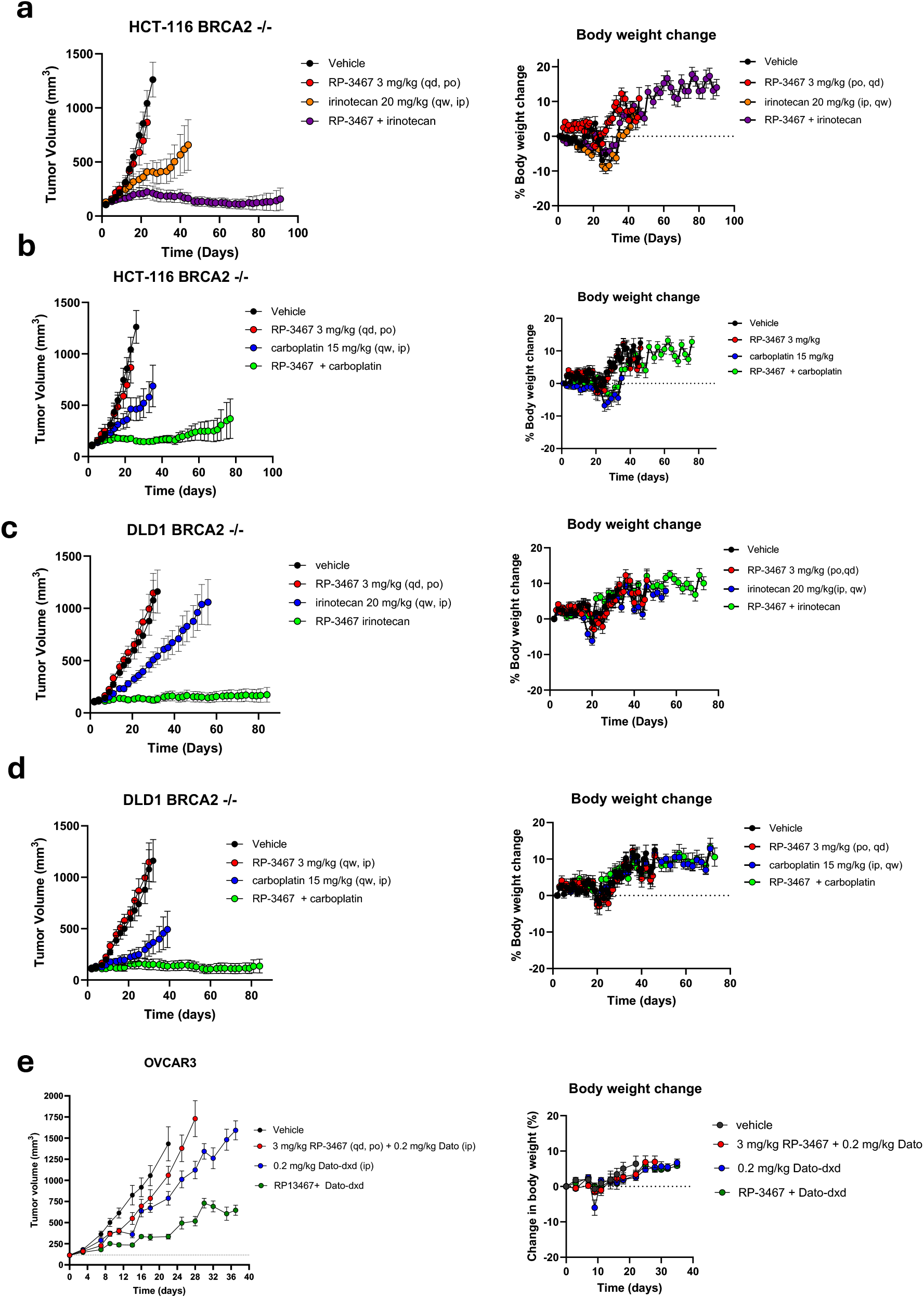
*In vivo* efficacy of RP-3467 in combination with chemotherapy or antibody-drug conjugates. Related to Figure 3. **a-f.** Tumor xenograft volume from HCT116 *BRCA2^-/-^* (a,b,c), DLD1 *BRCA2^-/-^* (d,e) and OVCAR3 (f) tumor bearing mice. Tumor volumes are mean +/- SEM; N=5-10 mice/group. Mice were treated orally with RP-3467 once daily for the duration of the experiments and (a) once daily via IP with 100 mg/kg Olaparib, (b-d) once weekly with 20 mg/kg irinotecan, (c,e) once weekly with 15 mg/kg carboplatin or (f) once on day 0 with either 2 mg/kg datopomab (Dato) or 2 mg/kg Datopomab-Deruxtecan (Dato-Dxd).

**Extended Data Figure 6.**
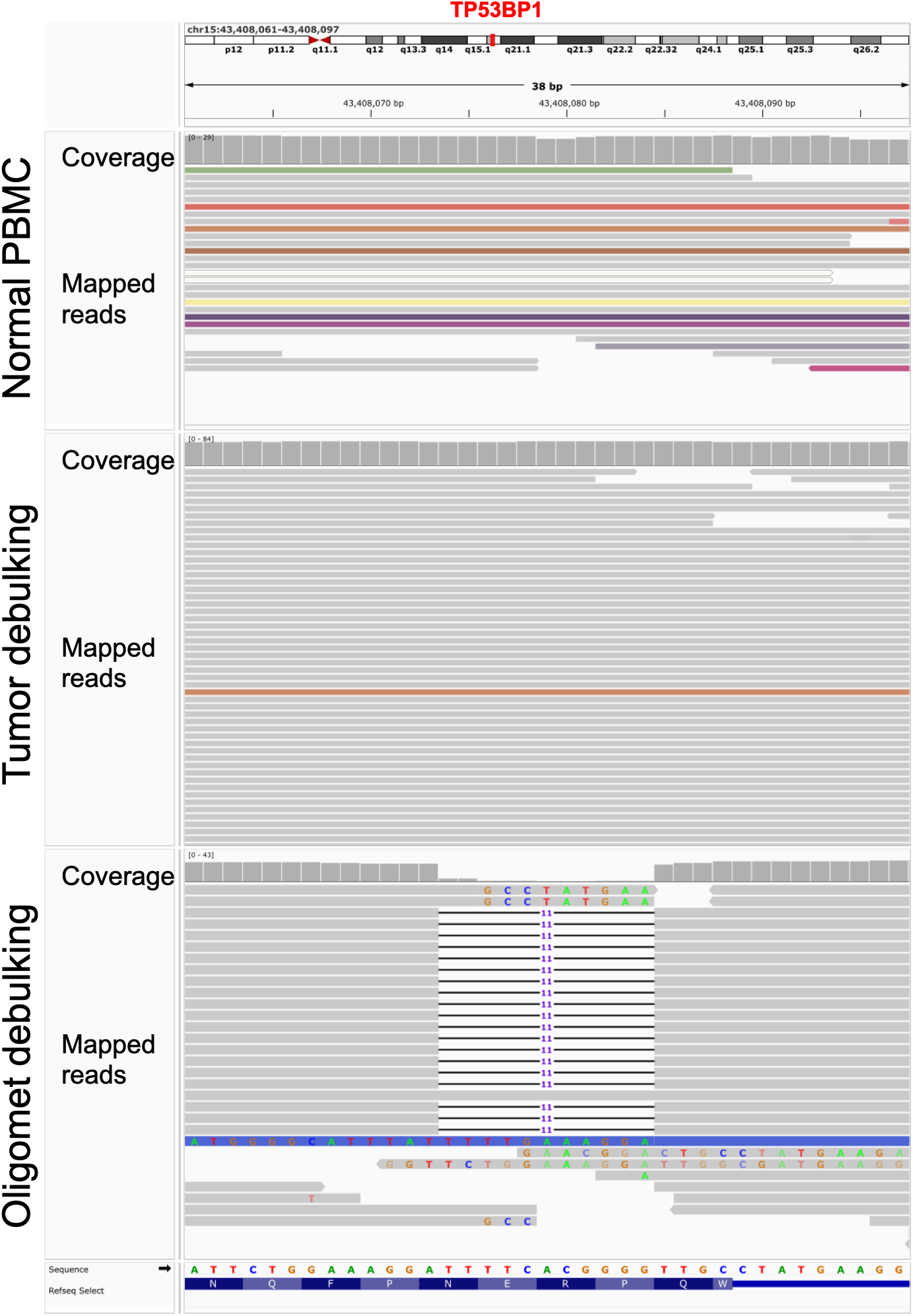
Genomic visualization of *TP53BP1 mutation*. Whole genome sequencing coverage and mapped reads for PBMC, tumor debulking, and oligomet debulking visualized in the Integrative Genomics Viewer. Visualization centered around the TP53BP1 locus where the 11 base pair deletion in the oligomet debulking sample is present.

**Extended Data Figure 7.**
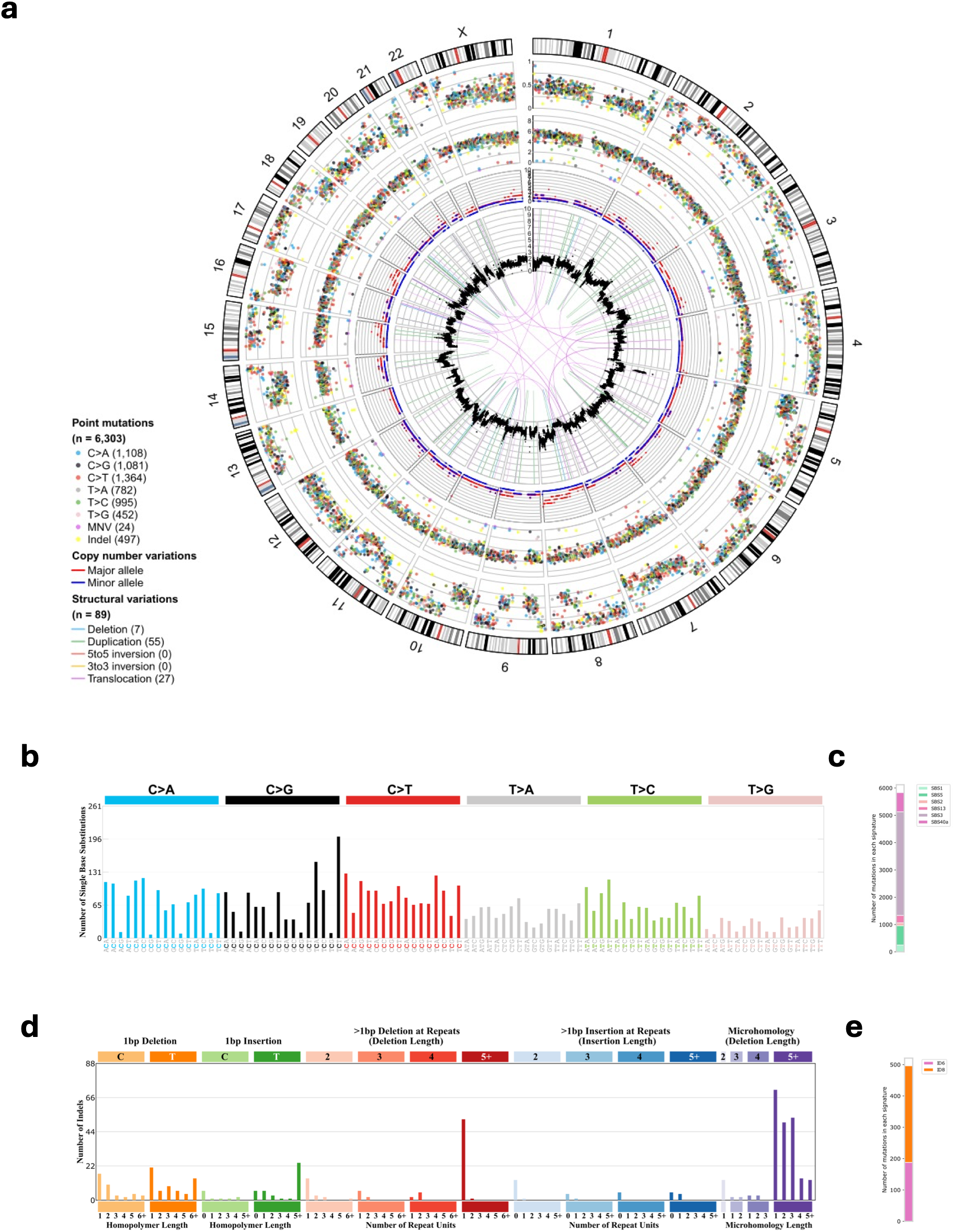
Whole genome sequencing analysis of patient samples. Related to Figure 6. Sequencing data from tumor debulking and oligomet debulking samples were compared to PBMC sequencing as described in methods. **a.** Circos plot indicating point mutations, copy number variations, and structural variations shared by tumor and oligomet debulking samples. **b.** Number of single-base substitution mutations organized into 96 genomic contexts according to preceding and succeeding base pair. **c.** Single-base substitution COSMIC mutational signature estimated composition of signal in (b). **d.** Number of insertions and deletions organized into 83 genomic contexts according to size, structure, and microhomology. **e.** Insertion-deletion COSMIC mutational signature estimated composition of signal in (d).

## MATERIALS AND METHODS

### Chemical compounds

RP-3467 was synthesized by Repare Therapeutics. Olaparib was purchased from Combi-Blocks. Pharmaron synthesized Saruparib. Carboplatin, SN-38, exatecan, Dato-DxD and SG3199 were purchased from MedChem Express. For *in vitro* experiments, all compounds except carboplatin were dissolved in DMSO to 10 mM stock concentrations and stored at -20°C for long-term storage. Carboplatin was made up in ddH_2_O / 0.3% (v/v) Tween-20 at 20 mM immediately before use. *In vivo* formulations are described below.

### Cell culture

HCT116 *BRCA2*^+/+^ and *BRCA2^-/-^* cells were purchased from ATCC (CCL-247) and ECACC General Cell Collection (18061301), respectively. They were grown in culture in McCoy’s media (Hyclone, SH30200.01) supplemented with 10% fetal bovine serum (FBS) and 1% Penicillin/Streptomycin. DLD1 *BRCA2^+/+^*and *BRCA2^-/-^* cells were purchased from DiscoverX (HD-PAR-086 and HD 105-007, respectively) and grown in RPMI media (Corning 10-040-CV) supplemented with 10% FBS, 1% Penicillin/Streptomycin and 2 mM sodium pyruvate (Corning 25-000-CI). DOTC24510 cells were purchased from ATCC (CRL-7920) and were grown in DMEM media (Corning 10-013-CV) supplemented with 10% FBS and 1% Penicillin/Streptomycin. MDAMB436 cells were purchased from ATCC (HTB-130) and cultured in DMEM + 10% FBS + 1% Penicilin/Streptomycin + 10 μg/ml insulin + 16 μg/ml gluthatione. OVCAR3 cells (ATCC) were cultured in RPMI-1640 + 20% FBS + 1% + 13 μg/ml insulin + 1% Penicilin/Streptomycin. DLD1 *POLQ^-/-^* GenScript generated cells by electroporating DLD1 cells with Cas9/sgRNA ribonucleoprotein complexes and selecting clones. The following sgRNA target sequence was used: 5’ AGGCAGCGACCAAGGCCGGG 3’. Knockout clones were validated by Sanger sequencing. All cell lines were maintained at 37°C and 5% CO_2_. All cells were routinely tested for mycoplasma and murine pathogens.

### Polθ ATPase enzymatic assay

The Polθ human ATPase domain (aa 1-894) was produced in *E. coli* as described in (*Mochirian, P. et al.,*^58^). 0.5nM enzyme was incubated with a 10-point concentration range of RP-3467 for 15 min at room temperature (RT) in the following buffer: 50 mM Tris Cl pH 7.5, 10% glycerol, 5 mM DTT, 10 mM MgCl_2_, 0.1 mg/ml BSA. 20 nM Fork C DNA substrate (see below) and 50 mM ATP were subsequently added, and the reaction was incubated at RT for 240 min. ATP consumption was measured using the ADP-Glo Kinase assay kit (Promega V6930) according to the manufacturer’s instructions. Luminescence was read on an Envision plate reader, and the IC_50_ value was determined in GraphPad PRISM by fitting the experimental data to a 4-parameter dose-response model.

Fork C was made by annealing the following DNA oligonucleotides:

Oligo 1: 5’-GCACTGGCCGTCGTTTTACGGTCGTGACTGGGAAAACCCTGGCG-3’

Oligo 2: 5’-TTTTTTTTTTTTTTTTTTTTTTCCAAGTAAAACGACGGCCAGTGC-3’

Oligo 3: 5’-TTGGAAAAAAAAAAAAAAAAAAAAAA-3’

Oligos were annealed by heating at 95°C for 5 min in a buffer containing 10 mM Tris-HCl pH7.5, 50 mM NaCl, 1 mM EDTA, and cooling to RT.

### HELQ, BLM, FANCM ATPase assays

DNA-dependent ATPase counter screen assays were performed in 384-well plates. ATP consumption was measured using the ADP-Glo Kinase assay kit (Promega V6930) according to the manufacturer’s instructions using either forked DNA duplex C (as per Polθ ATPase) or forked DNA duplex D substrates. Luminescence was read on an Envision plate reader and the IC50 value was determined in GraphPad PRISM by fitting the experimental data to a 4-parameter dose-response model. Specific conditions for each ATPase were as follows:

HELQ: 10nM purified recombinant HELQ (aa 1-1101) was incubated with compounds, 10 nM forked DNA duplex C, 100 μM ATP, 50 mM Tris Cl pH 7.5, 10% glycerol, 5 mM DTT, 10 mM MgCl_2_, 0.1 mg/ml BSA in a final volume of 15 μL at RT for 1 hr.

BLM: 1 nM purified recombination BLM (aa 636-1298) was incubated with compounds, 20 nM forked DNA duplex C, 100 μM ATP, 50mM Tris Cl pH 7.5, 10% glycerol, 5 mM DTT, 10 mM MgCl_2_, 0.1 mg/ml BSA in a final volume of 15 μL at RT for 1 hr.

FANCM: 6.3 nM purified recombinant FANCM (aa 73-645) was incubated with compounds, 2.5 nM forked DNA duplex D, 15 μM ATP in 25 mM Tris pH 7.0, 75mM NaCl, 5% glycerol, 0.005% NP40, 1 mM DTT, 0.5 mM MgCl_2_, 0.1 mg/ml BSA in a final volume of 15 μL at RT for 1 hr.

Fork D was made by annealing the following DNA oligonucleotides:

Oligo 1: 5’- GCACTGGCCGTCGTTTTACGGTCGTGACTGGGAAAACCCTGGCG -3’

Oligo 2: 5’- TTTTTTTTTTTTTTTTTTTTTTCCAAGTAAAACGACGGCCAGTGC -3’

Oligo 3: 5’- TTGGAAAAAAAAAAAAAAAAAAAAAA -3’

Oligo 4: 5’- CGCCAGGGTTTTCCCAGTCACGACC -3’

Oligos were annealed by heating at 95°C for 5 min in a buffer containing 10 mM Tris-HCl pH 7.5, 50 mM NaCl, 1mM EDTA, and cooling to RT.

### Cellular Thermal Shift Assay (CETSA)

CETSA experiments were performed as described by *Mochirian et al.*^58^. Briefly, an ePL-tagged FLAG-Polθ helicase domain (1-894) cloned into a pICP-ePL-C vector was stably expressed in K562 cells (ATCC, CCL-243). The cells were cultured in IMDM media containing 10% FBS, 1% Penicillin/Streptomycin, and geneticin. To perform CETSA, DMSO or RP-3467 was dispensed in a 96-well PCR plate with a Tecan D300E digital dispenser. 15×10^3^ cells were then added to each well and incubated for 1 hour at 37°C, followed by a heat pulse at 55°C for 3 min in a thermocycler. After the heat pulse, a 3-minute recovery step at 20°C was applied. The EFC working detection solution (lysis, complementation β-gal and substrate; InCELL Detection Kit, Eurofins DiscoverX, 96-0079) was added and incubated for 1 h in dark at room temperature. Chemiluminescence was read on an Envision luminescence plate reader. EC_50_ values were obtained by fitting the dose-response data to a four-parameter dose-response model in GraphPad PRISM.

### Cell proliferation assays

HCT116 *BRCA2^+/+^* (300 cells/well), HCT116 *BRCA2^-/-^*(600 cells/well), DLD1 *BRCA2^+/+^*(300 cells/well), DLD1 *BRCA2^-/-^* (600 cells/well), or DOTC24510 (1000 cells/well) cells were seeded in 96-well plates (Costar 3595) and incubated overnight in a tissue culture incubator (5% CO_2_, 37°C). The next day, compounds were added using a Tecan D300E dispenser. Final DMSO concentration was kept below 0.2%. Compound-containing media were replenished every 3 or 4 days. Cell confluence was monitored daily using the Incucyte® Live-Cell Analysis System until DMSO-treated wells reached 90% confluence (7 days for *BRCA2*^+/+^ and 12-14 days for *BRCA2^-/-^* cells; approximately six population doublings). Final confluence was determined using Incucyte® Cell-by-Cell Analysis Software Module, and IC_50_ values were calculated by fitting experimental data to a 4-parameter dose-response in GraphPad PRISM. For combination studies, ZIP synergy scores were computed using SynergyFinder^51^.

### Cas9-induced DNA double-strand break assay

Cas9/sgRNA ribonucleoprotein complexes were formed by co-incubating 40 µg of Alt-R® S.p. Cas9 Nuclease V3 (IDT, 1081059) with 240 pmol *AAVS1* sgRNA (5’ GGGGCCACTAGGGACAGGAT 3’) for 15 min at RT. Electroporation Enhancer (0.2 nmol; IDT, 1075916) was added at the end of the incubation period. HCT116 or DLD1 cells (1×10^6^) were nucleofected with the Cas9/sgRNA complex using a Lonza 4D nucleofector instrument (buffer SE / program EN113 for HCT116 and CM158 / SF for DLD1, respectively) and seeded in a 96-well plate (Costar 3595) at 35,000 cells/well. The DNA-PKcs inhibitor AZD7648 (Chemitek, CT-A7648; final concentration 0.5µM) and RP-3467 were then added using a Tecan D300E instrument and cells were incubated for 24 h. The next day, culture media were removed, and cells were lysed with 10 μl/well Sigma Extract-N-Amp Blood PCR kit lysis buffer (Millipore-Sigma, XNAB2) for 10 min at 75°C, followed by the addition of 90 μl/well of Sigma Extract-N-Amp Blood PCR kit neutralization buffer. A PCR mix to amplify the breakpoint site was prepared by combining 10.75 μl ddH_2_O, 5 μl Phusion HF buffer, 0.25 μl 50mM MgCl_2,_ 0.75 μl DMSO, 1.25 μl each of 5 μM F and R primers (5’ ACACTGACGACATGGTTCTACAAGTCTGTGCTAGCTCTTCCAGC 3’ and 5’ TACGGTAGCAGAGACTTGGTCTGTCACCAGATAAGGAATCTGCC 3’, respectively), 0.5 μl dNTP mix, 5 μl cell lysate, and 0.25 μl Phusion hot start flex DNA polymerase (New England Biolabs, M0535L). PCR was performed with the following conditions:

- 98°C, 30 s
- 15 cycles of:

`o` 98°C,10 s
`o` 72°C, 10s, -0.5°C/cycle
`o` 20°C, 20s
- 22 cycles of:

`o` 98°C,10 s
`o` 65°C, 10s
`o` 72°C, 20s
- 72°C, 3 min

PCR samples were Sanger-sequenced using the F primer and sequencing traces were analyzed with ICE (https://ice.editco.bio/#/)^52^ to quantify Polθ-mediated MMEJ repair outcomes (-5 and -12 bp indels; **Extended Data Figure 1**). Experimental data were fitted to a 4-parameter dose-response model in GraphPad PRISM to obtain IC_50_ values.

### Cell immunofluorescence

Cells were seeded on collagen-coated, black, clear-bottom 96-well imaging plates (Phenoplate 96, Revvity 6055700) and treated with serial dilutions of RP-3467 the next day, dispensed using a Tecan D300E instrument. Cells were cultured in the presence of the compound for 96 hours, after which the medium was removed, the cells were rinsed with PBS, and fixed with 4% paraformaldehyde (PFA) / PBS for 10 min at RT. Cells were washed 3x with PBS and permeabilized with 0.3% Triton X-100 / PBS for 30 min at RT. Cells were then blocked with PBG (0.2% w/v cold water fish gelatin / 0.5% w/v bovine serum albumin / PBS) for 30 min at RT, followed by incubation with primary antibodies diluted in PBG for 2 hours at RT. The following antibodies and dilutions were used: Rabbit anti-CIP2A, Novus NBP2-48710, 1:500; Mouse anti-H3pS10, Millipore-Sigma 05-806, 1:1000. Cells were washed 2-3 times with PBS and incubated with fluorescently labeled secondary antibodies (Alexa Fluor 555-conjugated goat anti-rabbit IgG, Invitrogen A-21428; Alexa Fluor 647-conjugated goat anti-mouse IgG A-21235) diluted at 1:1000 in PBG for 1 hour at RT. 0.5 mg/ml 4’,6-diamidino-2-phenylindole was included in the antibody mix to counterstain DNA. Finally, cells were washed 3x with PBS and imaged on an Operetta CLS or an Opera Phenix automated high-content microscope (Revvity) with a 40x water immersion objective in confocal mode. Detection of micronuclei and quantification of CIP2A staining intensity were performed using the Harmony software (Revvity) using built-in algorithms.

### Long-term MDAMB436 cell growth assay

On day one, 1x 10^5^ MDAMB436 cells were seeded in 6-well plates in the presence of DMSO, olaparib, RP-3467, or their combination as indicated in **Figure 2g**. When DMSO-treated cells reached ∼80% confluence (7-8 days), the cultures were collected by trypsinization, cells were counted, and the cells were reseeded on fresh plates as above. The procedure was repeated every 7-8 days until the assay was terminated on day 29.

### Rodent pharmacokinetic studies

All *in vivo* procedures were conducted at Repare Therapeutics (AdMare, Saint-Laurent, Canada), a CCAC (Canadian Council on Animal Care)-accredited vivarium, in accordance with the regulations and established guidelines of the AdMare Institutional Animal Care Committee. Female CD1 Nude or SCID-Beige mice (Charles River) were administered with test compound at a dose of 1 mg/kg intravenously using a solution formulation. Oral bioavailability was determined at a dose of 2.5 mg/kg using a solution formulation. The blood samples for the IV experiment were collected at pre-dose, 5, 15, 30 min, 1, 2, 4, 8, and 24 h time points. The blood samples for the PO experiment were collected at pre-dose, 0.5, 1, 2, 4, 6, 8, and 24 h time points. Pharmacokinetic studies in male CD rats were similarly performed but using an intravenous dose of 0.5 mg/kg and K3EDTA plasma collection. For drug-drug interaction studies, a cohort of mice (n=4), a satellite cohort to the efficacy studies, was treated with RP-3467 or olaparib alone or in combination to evaluate the pharmacokinetics of the doses used in the efficacy studies. Whole blood was collected pre-dose, and at time 0.5, 1, 2, 4, and 8 hrs post the morning dose; mice were treated again at 8 hours, and blood was collected at 10, 12, and 24 hours after 1 or 7 days of treatment. Micro-sampled whole blood was collected for mouse pharmacokinetic determinations using a previously described method^33^. All samples were quantified using a reversed-phase liquid chromatography gradient coupled to electrospray mass spectrometry operated in positive mode. PK parameters were calculated using non-compartmental analysis.

### Drug formulation

RP-3467 was prepared in a spray-dried powder form (50% with excipient) that was then suspended in 0.5% methylcellulose/sodium lauryl sulfate (0.5% MC / SLS) and was dosed via oral gavage daily (BID) at a dosing volume of 5 mL/kg in 0.5% methylcellulose + 0.02% sodium lauryl sulfate. For efficacy studies, olaparib was dosed orally by gavage or intraperitoneally daily (QD) at 5 mL/kg in 10% DMSO/90% (30% HP-β-CD in water). Olaparib was suspended in 10% DMSO / 90% (10% HP-β-CD in PBS). Saruparib in 0.5% methylcellulose / 0.02% SLS. Carboplatin was dissolved in water, irinotecan in 5%DMSO in PBS, and Dato-DXd was diluted in PBS.

For hematological toxicity assessments, olaparib was formulated in chow to reflect clinically relevant exposure better and administered continuously to female CD-1 mice (6-9 weeks). To formulate olaparib in chow, pre-weighed olaparib was dissolved in acetone and then added to a pre-weighed amount of rodent chow (Teklad T.2918M.15) while mixing a Kitchen Aid Stand Mixer 6QT 575Watt for 1 hour. The formulated chow was then transferred into a bottle and placed under vacuum for 2 hours to complete the drying process. The concentration of olaparib in each batch of formulated chow was verified to be within 10% of the desired concentration by HPLC.

### Cell line and patient-derived xenografts

In vivo experiments were conducted at Repare Therapeutics in a Canadian Council on Animal Care-accredited vivarium, in accordance with Institutional Animal Care Committee-approved protocols. HCT116 *BRCA2*^+/+^, HCT116 *BRCA2 ^-/-^*, MDA-MB-436, and OVCAR3 cells were implanted subcutaneously (at 1 × 10^7^ cells/mouse) into the flanks of female SCID-Beige mice (6–8 weeks old; Charles River) in a suspension of 1:1 media:Matrigel (Matrigel Corning; CB35248). Patient-derived xenograft studies were conducted at Xentech (France) and CrownBio (China). PDX fragments of ∼ 20 mm^³^ were implanted from donor mice into 5- to 9-week-old female athymic nude mice after tumor expansion *in vivo*. RP-3467 was dosed via oral gavage daily (QD). Clinical signs and body weight were monitored three times per week. Tumor growth was monitored using an electronic caliper, and tumor volume (TV) was calculated using the formula: TV = 0.52 × L × W^^2,^ where L and W are the tumor lengths and widths, respectively. Body weight change is represented as a change in BW using the formula: %BW change = (BW_last_-BW_day0_) / BW_day0_ x 100.

### Complete blood counts

Whole blood was collected from anesthetized mice into K_3_-EDTA BD Microtainer MAP Microtubes (Thermo Fisher, 22-253-270) by cardiac puncture. Hematology analysis was performed with a Sysmex XN-1000TM hematology analyzer (Sysmex America Inc.) according to the manufacturer’s instructions.

### Analysis of micronuclei in FFPE tissues

Tumor xenograft tissues were stained with DAPI, and slides were scanned on either an Akoya Polaris or PhenoImager HT slide scanners (Akoya Biosciences). Digital images were analyzed using HALO® AI software (Indica Labs). To detect micronuclei, nuclei and their corresponding DAPI signals were first segmented to generate a nuclear mask. The DAPI image was then duplicated and, using the nuclear mask, separated into two channels: one containing only nuclear signal and the other containing extra-nuclear DAPI signal. The HALO® FISH-IF module was adapted for automated detection, and micronuclei were defined as punctate extra-nuclear DAPI signals located near a tumor cell nucleus. Necrotic regions and non-tumor cells within the xenograft tissue were excluded from the analysis. If a micronucleus was located near two nuclei, it was assigned to the nearest cell based on the centroid position of the nuclear mask.

### CIP2A immunofluorescence on FFPE tissues

FFPE tissue sections were deparaffinated in HistoClear II (Electron Microscopy Sciences, 64111) twice, for 10 and 5 min, followed by 5 min in 100% ethanol. Sections were then rehydrated through a graded ethanol series (95%, 80%, 70%, and 50%) for 2 min each. After a PBS wash, slides were submerged in Borg Decloaker buffer (Biocare Medical, BD1000), and an antigen retrieval step was performed in a pressure cooker (Biocare Medical, DC2012) at 110°C for 30 min. After cooling, slides were incubated in blocking buffer (5% normal goat serum, 1% glycerol, 0.1% BSA, 0.1% Fish skin gelatin [Sigma G-7765] in PBS) for 1 hour at RT.

Primary antibodies were diluted in blocking buffer: 1:200 for CIP2A (Novus Biologicals, NBP2-48710) and 1:1000 for phospho-Histone H3 (Ser10, clone 3H10) (Millipore, 05-806). Sections were incubated with primary antibodies overnight at 4°C.

Following washes, secondary antibodies were applied for 1 hour at RT: 1:200 anti-rabbit Alexa Fluor 647 and 1:1000 anti-mouse Alexa Fluor 555. Coverslips were mounted using ProLong Gold Antifade Mountant (Invitrogen, P36931). Imaging was performed using Opera Phenix Plus high-content screening system (Revvity) in PreciScan mode. Image analysis was carried out using Harmony software (Revvity).

### Clinical study design and treatment

The POLAR study is a multicenter, open-label Phase I trial to investigate the safety, PK, pharmacodynamics, and preliminary efficacy of the Polθ inhibitor RP-3467 alone or in combination with the poly-ADP ribose polymerase (PARP) inhibitor (PARPi) olaparib in adults with molecularly selected advanced solid tumors. The trial is enrolling participants with breast, ovarian, prostate, or pancreatic cancers that harbor a deleterious alteration in *BRCA1*, *BRCA2*, *PALB2*, *RAD51B/C/D,* or that have homologous recombination deficiency (HRD+). The study was conducted in accordance with the Declaration of Helsinki and Council for International Organizations of Medical Sciences International Ethical Guidelines, applicable International Conference on Harmonization Good Clinical Practice Guidelines, and applicable laws and regulations. All patients provided written informed consent to adhere to the clinical protocol and provide serial blood samples and tumor tissue. The protocol was approved by the institutional review board or ethics committee at each participating institution. Clinical data presented in this manuscript were cut with an end date of July 31, 2025.

### ctDNA analysis

Blood for retrospective analysis was collected in two 10 mL K_2_-EDTA tubes pre-treatment and throughout treatment. Plasma and buffy coats were isolated immediately following phlebotomy by centrifugation, frozen at -80°C, and bio-banked. Plasma and buffy coat aliquots were processed and analyzed in batches at Guardant Health. Upon receipt of frozen plasma, cell-free DNA (cfDNA) was extracted and sequenced at Guardant Health, the Guardant Infinity^TM^ platform (Guardant Health, Inc., Redwood City, CA)

### Whole Genome Sequencing and analyses

Genomic DNA from one normal PBMC sample and two tumor FFPE samples was prepared for sequencing using the Watchmaker DNA Library Preparation Kit (Watchmaker Genomics, Boulder, CO). Samples were sequenced on an Illumina NovaSeq X Plus (San Diego, CA) to 25x (PBMC) and 50x (tumor) depth, as previously described^53^. Data analysis, performed with the Inocras CancerVision system (Inocras Inc., San Diego, CA), included mapping to GRCh38.p14 using bwa-mem2^54^ and somatic mutations calling using Strelka2 v2.9.10^55^ and Mutect2-GATK v4.2.0.0^56^. Structural variant analysis was carried out using Manta v1.6.0^57^. Only variants with a PASS quality control filter option in both tumor samples were used in the analysis. Structural variants with 5 or fewer spanning paired-read or split read support were not included in the analysis. Structural variants that could not be accurately classified as deletion, inversion, tandem duplication or translocation were removed from the analysis. Tumor ploidy estimation, segmentation, and copy number analysis was carried out as previously described^53^. Homologous recombination deficiency was assessed using CHORD^47^.

## Data availability

Raw sequencing data for AAVS1-Seq will be deposited in NCBI’s Sequence Read Archive (SRA) under BioProject. Requests for materials and/or questions regarding any of the constructs, cell lines, or other data described should be addressed to the corresponding authors. Source data are provided in this paper, and raw data will be available in Mendeley.

